# The impact of anorexia nervosa and BMI polygenic risk on childhood growth: a 20-year longitudinal population-based study

**DOI:** 10.1101/2020.10.15.20200600

**Authors:** Mohamed Abdulkadir, Christopher Hübel, Moritz Herle, Ruth J.F. Loos, Gerome Breen, Cynthia M. Bulik, Nadia Micali

**Affiliations:** Department of Pediatrics Gynaecology and Obstetrics, Faculty of Medicine, University of Geneva, Geneva, Switzerland; Department of Psychiatry, Faculty of Medicine, University of Geneva, Geneva, Switzerland; Social, Genetic & Developmental Psychiatry Centre, Institute of Psychiatry, Psychology & Neuroscience, King’s College London, UK; UK National Institute for Health Research (NIHR) Biomedical Research Centre for Mental Health, South London and Maudsley Hospital, London, UK; National Centre for Register-based Research, Aarhus Business and Social Sciences, Aarhus University, Aarhus, Denmark; Department of Medical Epidemiology and Biostatistics, Karolinska Institutet, Stockholm, Sweden; Great Ormond Street Institute of Child Health, University College London, London, UK; Department of Biostatistics & Health Informatics, Institute of Psychiatry, Psychology & Neuroscience, King’s College London, UK; Novo Nordisk Foundation Center for Basic Metabolic Research, Department of Health and Medical Sciences, University of Copenhagen, Copenhagen, Denmark; Icahn School of Medicine at Mount Sinai, New York, New York, USA; Department of Psychiatry, University of North Carolina at Chapel Hill, Chapel Hill, NC, USA; Department of Nutrition, University of North Carolina at Chapel Hill, Chapel Hill, NC, USA

**Author notes:** contributed equally to this work (Mohamed Abdulkadir, Christopher Hübel, Moritz Herle, Cynthia M. Bulik, Nadia Micali). Correspondence to: Dr. Micali, Department of Psychiatry, Faculty of Medicine, University of Geneva, Geneva, Switzerland.

**Keywords:** Eating disorder, Avon Longitudinal Study of Parents and Children (ALSPAC), growth trajectories, body mass index, fat mass index, fat-free mass index, bone mineral density, lean mass index

## Abstract

**Background:** Deviating growth from the norm during childhood has been associated with anorexia nervosa (AN) and obesity later in life. In this study, we examined whether polygenic scores (PGSs) for AN and BMI are associated with growth trajectories spanning the first two decades of life.

**Methods:** AN-PGS and BMI-PGS were calculated for participants of the Avon Longitudinal Study of Parents and Children (ALSPAC; N=8,654). Using generalized (mixed) linear models, we associated PGSs with trajectories of weight, height, body mass index (BMI), fat mass index (FMI), lean mass index (LMI), and bone mineral density (BMD).

**Results:** Female participants with one SD higher AN-PGS had on average 0.004% slower growth in BMI between the ages 6.5-24 years and a 0.4% slower growth in BMD between the ages 10-24 years. Higher BMI-PGS was associated with faster growth for BMI, FMI, LMI, BMD, and weight trajectories in both sexes throughout childhood. Female participants with both a high AN-PGS and a low BMI-PGS showed slower growth compared to those with both a low AN-PGS and a low BMI-PGS.

**Conclusion:** AN-PGS and BMI-PGS have detectable sex-specific effects on growth trajectories. Female participants with high AN-PGS and low BMI-PGS likely constitute a high-risk group for AN as their growth was slower compared to their peers with high PGS on both traits. Further research is needed to better understand how the AN-PGS and the BMI-PGS co-influence growth during childhood and whether high BMI-PGSs can mitigate the effects of a high AN-PGS.

## Introduction

Anorexia nervosa (AN) is a serious psychiatric disorder that is characterized by low fat and lean mass [1–3]. Observations from genome-wide association studies (GWASs) suggest that genomic variants that influence body composition are also associated with psychiatric traits [4]. Genetically, AN is negatively correlated with body mass index (BMI), fat mass, fat-free mass, and obesity [4], suggesting that biological mechanisms contributing to AN may also influence body composition. This association is supported by several studies showing that low premorbid BMI is associated with AN in adolescence [5,6]. Furthermore, an Avon Longitudinal Study of Parents and Children (ALSPAC) study reported that individuals who go on to develop AN followed lower BMI trajectories (as early as age 2 years) compared to their peers that did not develop an eating disorder (ED) [7].

In contrast to low body weight, high body weight has not only been associated with increased risk for cardiovascular disease but also to psychiatric disorders (e.g., mood disorders and anxiety disorders) [8,9]. In addition, individuals with high body weight face stigmatization and discrimination from the public and health professionals, which can exacerbate the negative health effects of obesity [10–12]. Similar to AN, BMI has been extensively studied on a genetic level and is a heritable polygenic trait [13,14]. Khera et al. [15] reported that a BMI polygenic score (BMI-PGS), calculated by summing the BMI-increasing alleles of all variants of a BMI GWAS, weighted by their reported effect sizes [13], is associated with body weight at different timepoints during childhood and adolescence. For example, individuals with a BMI-PGS in the top decile have a modest, yet significant, higher birth weight (+60 grams) than individuals with an BMI-PGS in the bottom decile [15]. However, this difference in weight increases over time, reaching 3.5 kg by age 8 years and 12.3 kg by age 18 years [15]. These findings highlight differences in growth associated with the polygenic liability to high BMI.

In summary, both AN and BMI have a genetic component that can be summarized by PGSs, and these genetic components are inversely correlated. However, it is unclear how genetic risk for both traits, individually and combined, affect growth developmentally during the first two decades of life. We identified individuals considered to be at high risk (defined as PGS in the top two deciles) for either AN or BMI and compared them with their peers with lower risk (defined as PGS in the lower 8 deciles) for the same trait. We then studied the longitudinal effects of the AN-PGS [4] and the BMI-PGS [14] (separately and combined) on weight, height, BMI, fat mass index (FMI), lean mass index (LMI), and bone mineral density (BMD) growth trajectories during the first two decades of life using data from ALSPAC [16– 20]. We hypothesized that a higher AN-PGS would be associated with slower growth for the weight, BMI, FMI, LMI, and BMD trajectories. Previous studies reported no genetic correlation between height and AN and therefore we used a height trajectory as a negative control (3). We also hypothesized that a higher BMI-PGS would be associated with faster growth trajectories. Lastly, we hypothesized that individuals with both a high AN-PGS and low BMI-PGS would represent a subgroup at higher risk for poor growth (slower growth) compared to those with both a low AN-PGS and a low BMI-PGS.

## Methods

### Participants

The ALSPAC study is an ongoing population-based birth cohort study of 14,541 mothers and their children (that were born between 1 April 1991 and 31 December 1992) residing in the southwest of England (UK) [16–20]. From the 15,541 pregnancies, 13,988 were alive at 1 year. At age 7 years this sample was bolstered with an additional 913 children. The total sample size for analyses using any data collected after the age of seven is therefore 15,454 pregnancies; of these 14,901 were alive at 1 year of age. Participants are assessed at regular intervals using clinical interviews, self-report questionnaires, medical records, and physical examinations. Study data were collected and managed using REDCap electronic data capture tools hosted at University of Bristol [21,22]. REDCap (Research Electronic Data Capture) is a secure, web-based software platform designed to support data capture for research studies, providing 1) an intuitive interface for validated data capture; 2) audit trails for tracking data manipulation and export procedures; 3) automated export procedures for seamless data downloads to common statistical packages; and 4) procedures for data integration and interoperability with external sources. Please, note that the study website contains details of all the data that are available through a fully searchable data dictionary and variable search tool and reference the following webpage: http://www.bristol.ac.uk/alspac/researchers/our-data/. To avoid potential confounding due to relatedness, one sibling per set of multiple births was randomly selected to guarantee independence of participants - this resulted in the removal of 75 individuals. Furthermore, individuals who are closely related to each other defined as a phi hat > 0.2 (calculated using PLINK v1.90b) were removed; specifically, we removed any duplicates or monozygotic twins, first-degree relatives (parent-offspring and full siblings), and second-degree relatives (half-siblings, uncles, aunts, grandparents, and double cousins).. The authors assert that all procedures contributing to this work comply with the ethical standards of the relevant national and institutional committees on human experimentation and with the Helsinki Declaration of 1975, as revised in 2008. Ethical approval for the study was obtained from the ALSPAC Ethics and Law Committee and the Local Research Ethics Committees (Bristol and Weston Health Authority: E1808 Children of the Nineties: ALSPAC, 28th November 1989 (for details see: www.bristol.ac.uk/alspac/researchers/research-ethics/). Informed consent for the use of data collected via questionnaires and clinics was obtained from participants following the recommendations of the ALSPAC Ethics and Law Committee at the time. The main caregiver initially provided consent for child participation and from the age 16 years the offspring themselves have provided informed written consent.

### Measures

*Weight, height, and body mass index (BMI)*. Numerous measurements of weight and height were collected from different sources (i.e., routine clinic visits, information collected from midwives, linkage to child health records) between birth and age 24 years. Information on weight was collected at research clinic visits annually up to the age 14 years and further clinic measurements at the ages 16, 18, and 24 years using the Tanita Body Fat Analyzer (Tanita TBFUK Ltd.) to the nearest 50 grams. During the same clinic visits, height (standing) was measured to the nearest millimeter with shoes and socks removed using a Holtain stadiometer (Holtain Ltd, Crymych, Pembs, UK). The different measurements of weight and height were highly correlated (**Figure S1**) across various methods [16,23]. Information on child and adolescent BMI (weight in kilograms / height squared in meters) was derived using weight and height measurement obtained during clinic visits.

#### Fat mass index, lean mass index, and bone mineral density (BMD)

All ALSPAC participants were invited to undergo a whole-body dual-energy x-ray absorptiometry (DXA) scans using the Lunar Prodigy dual-energy X-ray absorptiometry scanner as part of face-to-face visits at the ages of 10, 12, 14, 16, 18, and 24 years. Fat mass index was calculated by dividing total body fat mass (in kilograms) by height (in meters) squared. Similarly, lean mass index was calculated by dividing total lean mass divided by height (in meters) squared. Additionally, whole-body (minus head) BMD was also estimated using the Lunar Prodigy dual-energy X-ray absorptiometry scanner.

### Trajectory modeling

#### Censoring for the presence of an eating disorder

To derive the trajectories for each outcome, we censored for the presence of any eating disorder (ED); i.e., AN, bulimia nervosa, and binge-eating disorder. Information on a probable ED was available at age 14, 16, and 18 years (see [24,25] for more information on how ED diagnoses were derived). The presence of an ED diagnosis at age 14 years meant that all values for that individual regarding their measurement (BMI, FMI, LMI, etc.) at age 14 years up to age 24 years were set to missing. This was also done for the presence of an ED diagnosis at age 16 years (set values at age 16 and beyond to missing), and 18 years (set values at age 18 and beyond to missing). Therefore, censoring did not lead to loss of participants in the analyses, but rather loss of observations (N = 1,055). This censoring allowed us to derive unbiased results in the following longitudinal modelling, while retaining the largest amount of data possible. This is important, as these models are sensitive to outliers and including individuals with EDs would likely introduce extreme values in the distribution.

#### Spline modeling (*Table S1 and Figure S2*)

To capture the potential impact of AN on growth, we derived a BMI trajectory across both childhood and adolescence. Prior to analyses, BMI values (**Table S1** and **Figure S2**) were transformed using the natural log due to the right-skewed distribution of the data. Spline models involve placing spline points (knots) at time points where the direction of growth changes. This is necessary as children’s growth in the first two decades of life is not linear, but follows a more complex pattern, rendering standard growth models unsuitable to accurately reflect the data [26]. The advantage of linear spline models is that they allow knot points to be fitted at different ages to derive periods of change (between the knots) that are approximately linear. After visual inspection of the BMI medians at each time point, two spline points (knots), in addition to the starting point (intercept) at 4 months and the final data wave (24 years), were placed, creating the following periods of linear growth between 4 months to age 1 year, 1 year to 6.5 years, and age 6.5 and 24 years. Mixed effects spline models were run to describe the longitudinal growth outcomes. The mixed effects framework lends itself to the analyses of repeated measures as it accounts for the non-independence of measures within an individual. After fitting model fitting, we extracted parameters of slopes using the best linear unbiased predictions (BLUPs). The linear spline modeling resulted in three slopes (coefficients), which correspond with the slopes in the periods of growth between age 4 months and 1 year, growth between age 1 year and 6.5 years, and growth between age 6.5 year and 24 years. Spline models were obtained using STATA (v 15).

### Genotyping

Genotype data were available for 9,915 out of the total of 15,247 ALSPAC participants. Participants were genome-wide genotyped on the Illumina HumanHap550 quad chip. Following quality control of the genetic data, a total of 8,654 participants with genotyping data and at least one outcome measure were included in the analyses (**Table 1**). Details of the quality control checks are described in the **Supplementary Information**.

**Table 1:** Descriptive data from the Avon Longitudinal Study of Parents and Children (ALSPAC). Data presented on age, body mass index (BMI), fat mass index (FMI), lean mass index (LMI), weight, and height^a^. The sample size per outcome varies.

|  |  | Female |  |  | Male |
| --- | --- | --- | --- | --- | --- |
| Body composition measure | Age (years) | N | Median (IQR) | N | Median (IQR) |
| BMI (kg/m<sup>2</sup>)<sup>b</sup> | 10 | 3,072 | 17.31 (15.77, 19.41) | 3,026 | 16.77 (15.61, 18.73) |
|  | 12 | 2,902 | 18.6 (16.74, 21.14) | 2,797 | 17.96 (16.43, 20.5) |
|  | 14 | 2,433 | 20.17 (18.43, 22.58) | 2,394 | 19.28 (17.71, 21.45) |
|  | 16 | 1,496 | 20.95 (19.47, 23.14) | 1,289 | 20.83 (19.14, 22.78) |
|  | 18 | 1,986 | 22.14 (20.35, 24.78) | 1,681 | 21.76 (20.08, 24.34) |
|  | 24 | 1,671 | 23.63 (21.47, 27.04) | 1,131 | 24.25 (21.97, 27.09) |
| FMI (kg/m<sup>2</sup>)<sup>c</sup> | 10 | 2,932 | 4.4 (3.14, 6.18) | 2,880 | 2.96 (2.08, 4.67) |
|  | 12 | 2,862 | 4.91 (3.52, 7.11) | 2,750 | 3.65 (2.53, 5.9) |
|  | 14 | 2,406 | 5.77 (4.28, 7.74) | 2,353 | 3.11 (2.09, 5.26) |
|  | 16 | 1,266 | 6.5 (5.00, 8.36) | 1,066 | 2.67 (1.83, 4.14) |
|  | 18 | 1,895 | 7.13 (5.68, 9.28) | 1,624 | 3.37 (2.22, 5.64) |
|  | 24 | 1,618 | 8.07 (6.43, 10.61) | 1,100 | 5.7 (4.32, 7.8) |
| LMI (kg/m<sup>2</sup>)<sup>c</sup> | 10 | 2,932 | 12.07 (11.5, 12.69) | 2,880 | 12.98 (12.43, 13.55) |
|  | 12 | 2,862 | 12.64 (11.95, 13.42) | 2,750 | 13.28 (12.63, 14) |
|  | 14 | 2,406 | 13.39 (12.7, 14.1) | 2,353 | 14.87 (13.9, 15.92) |
|  | 16 | 1,266 | 13.56 (12.81, 14.3) | 1,066 | 15.93 (14.76, 16.98) |
|  | 18 | 1,895 | 13.86 (13.17, 14.64) | 1,624 | 17.19 (16.16, 18.18) |
|  | 24 | 1,618 | 14.8 (13.95, 15.82) | 1,100 | 17.45 (16.25, 18.83) |
| BMD (g/cm<sup>2</sup>)<sup>c</sup> | 10 | 2,965 | 0.77 (0.74, 0.81) | 2,900 | 0.78 (0.75, 0.82) |
|  | 12 | 2,865 | 0.85 (0.8, 0.9) | 2,756 | 0.84 (0.80, 0.89) |
|  | 14 | 2,406 | 0.96 (0.91, 1.01) | 2,354 | 0.95 (0.90, 1.01) |
|  | 16 | 2,050 | 1 (0.96, 1.05) | 1,952 | 1.06 (0.99, 1.12) |
|  | 18 | 1,902 | 1.04 (0.99, 1.09) | 1,634 | 1.14 (1.08, 1.21) |
|  | 24 | 1,619 | 1.19 (1.13, 1.26) | 1,101 | 1.32 (1.23, 1.39) |
| Weight (kg)<sup>b</sup> | 10 | 3,105 | 33.6 (29.6, 38.8) | 3,045 | 33 (29.4, 37.8) |
|  | 12 | 2,904 | 42.8 (37, 50.2) | 2,801 | 40.6 (35.8, 47.6) |
|  | 14 | 2,433 | 53.4 (47.8, 60.2) | 2,394 | 53 (46.8, 61.35) |
|  | 16 | 1,599 | 57 (52, 64) | 1,330 | 66 (59, 74) |
|  | 18 | 1,986 | 61 (55.1, 68.4) | 1,683 | 70.4 (63.7, 79) |
|  | 24 | 1,671 | 64.9 (58.7, 75.55) | 1,131 | 79 (70.7, 88.5) |
| **Height (cm) <sup>b</sup>** | 10 | 3,074 | 139.05 (135, 143.4) | 3,027 | 139.8 (135.7, 143.9) |
|  | 12 | 2,904 | 151.4 (146.4, 156.2) | 2,797 | 149.8 (145.1, 154.7) |
|  | 14 | 2,438 | 162.1 (157.8, 166.3) | 2,394 | 165.2 (158.9, 170.8) |
|  | 16 | 1,666 | 165 (160, 170) | 1,361 | 178 (173, 183) |
|  | 18 | 1,988 | 165.1 (161.1, 169.2) | 1,683 | 179 (174.45, 183.4) |
|  | 24 | 1,672 | 165.8 (161.88, 170) | 1,131 | 180 (175.5, 184.5) |

IQR = interquartile range; BMI, body mass index (weight in kilograms/height<sup>2</sup> in meters); FMI, fat mass index (fat mass in kilograms/height<sup>2</sup> in meters); LMI, lean mass index (lean mass in kilograms/height<sup>2</sup> in meters); BMD, bone mineral density.
<sup>a</sup> Observations were censored for the presence of an eating disorder (ED); i.e., anorexia nervosa, bulimia nervosa, and binge-eating disorder. Information on a probable ED was available at age 14, 16, and 18 years [24,25]. The presence of an ED diagnosis at age 14 years meant that all values for that individual regarding their measurement (BMI, FMI, LMI, etc.) at age 14 years up to age 24 years were set to missing. This was also done for the presence of an ED diagnosis at age 16 years (set values at age 16 and beyond to missing), and 18 years (set values at age 18 and beyond to missing).
<sup>b</sup> BMI was calculated using objectively measured weight and height during a routine clinic visit at age 24 years. Height was measured to the nearest millimeter using a Harpenden Stadiometer (Holtain Ltd.) and weight was measured using the Tanita Body Fat analyzer (Tanita TBF UK Ltd.) to the nearest 50 grams. To limit the size of this table only the BMI values at age 10 years and later are shown. For BMI values prior to age 10 years see Table S1.
<sup>c</sup> Fat mass, lean mass, and bone mineral density (BMD) were derived using a Lunar Prodigy dual emission x ray absorptiometry (DEXA) scanner (GE Medical Systems Lunar, Madison, WI, USA). FMI and LMI were calculated by dividing each measure (in kilograms) by height<sup>2</sup> (in meters). Bone mineral density (BMD) was calculated for the whole body excluding the head values.

### PGS calculations

The AN-PGS was derived from the GWAS on AN by the Eating Disorders Working Group of the Psychiatric Genomics Consortium (PGC-ED; n_cases_ = 16,992, n_controls_ = 55,525) [4] and the BMI-PGS was derived from the BMI GWAS conducted by the Genetic Investigation of ANthropometric Traits (GIANT; n ∼700,000 individuals) [14]. The datasets will be referred to as the AN and BMI discovery cohorts, respectively. The PGSs were calculated using the PRS-CS software; a method that infers posterior SNP effect sizes under continuous shrinkage (CS) priors [27].

Furthermore, we derived a categorical variable using the AN-PGS and BMI-PGS as following; we dichotomized the AN and the BMI PGS based on a cut-off value at the 8^th^ decile of the PGS distribution. This value was selected based on previous studies that reported that individuals in top deciles of BMI-PGS and schizophrenia-PGS are at greater risk than those in lower deciles for being overweight and being diagnosed with schizophrenia, respectively [15,28]. We did not choose the highest decile as cut-off as this would have resulted in a small sample size and thus low statistical power for this particular bin. Individuals with a PGS lower than the 8^th^ decile were grouped into a “low PGS” group, while those with a PGS score at or higher than 8^th^ decile were grouped into a “high PGS” group. Based on this grouping, we were able to determine four categories; individuals with a (1) low PGS for AN and low PGS for BMI, (2) high PGS for AN and low PGS for BMI, (3) low PGS for AN and high PGS for BMI, and (4) high PGS for AN and high PGS for BMI. The “low AN-PGS and low BMI-PGS” group was used as the reference category in the analyses.

### Statistical analyses

#### Univariate polygenic score analyses

In the first set of analyses, we addressed whether the AN-PGS and BMI-PGS separately were associated with body composition trajectories. For the BMI spline trajectory, we regressed each derived slope onto the standardized AN-PGS or the BMI-PGS and the first four ancestry informative principal components. Data for FMI, LMI, and BMD were measured objectively at 10, 12, 14, 16, 18, and 24 years during face-to-face visits, as described above. For uniformity, we restricted analyses of weight and height to measurements at these ages as growth is more linear from mid-childhood onwards. Height was included to test the quality of methodological approach as a negative control for the association with the AN-PGS. (3). FMI, LMI, BMD, weight, and height trajectories were analyzed using linear mixed effects regression (LMER) using the LMER function from the LME4 package in R [29]; the standardized AN-PGS or BMI-PGS, the first four ancestry-informative principal components, and age were added as fixed effects. For the linear mixed effects models, the intercept and the slope were all allowed to vary randomly across individuals. We stratified our analyses on biological sex; phenotypic sex differences in body composition have been reported in the general population that are detectable as early as adolescence [30,31]. Considering that the BMI slopes are highly correlated, we additionally corrected for slope(s) that preceded the one that is being analyzed; i.e., for the model in which the slope between the ages 1 year and 6.5 years is the outcome we controlled for the slope preceding this period (slope between the ages 4 months and 1 year) and for the for the model in which the slope between the ages 6.5 year and 24 years is the outcome we controlled for the slopes preceding this period (slope between the ages 4 months and 1 year and the slope between ages 1 year and 6.5 years). We report for each model the beta (as a measure of effect size, with 95% confidence intervals), and the percent change in the estimated parameter to ease the interpretation of the beta point estimates.

Correction for multiple testing across all tests (N = 648) was done by calculating False Discovery Rate-corrected *Q*-values [32]. The significance threshold was met if the False Discovery Rate-adjusted *Q* was < 0.05.

#### Extreme group comparison of the polygenic score analyses

The effects of PGS are often most discernable in the most extreme deciles. We therefore tested whether groups characterized by a particularly high PGS load differ from one another. Note that this differs from testing a formal interaction between the AN-PGS and the BMI-PGS, as our focus is on understanding the difference in growth in the extreme end of the PGS distribution. In this second set of analyses, we used LMER to determine the association between the body composition measures (BMI, FMI, LMI, BMD, weight, and height) and the derived categorical AN and BMI PGS. For each regression model, the derived categorical variable of the AN-PGS or BMI-PGS, age, and the first four ancestry informative principal components were included as fixed effects. We included random intercepts and slope for each individual in the model to account for variance in body composition measures that is due to inter-individual differences. The analyses were stratified by sex, given differences in body composition [31]. The “low AN-PGS and low BMI-PGS” group was used as the reference category in the analyses. For this set of tests, correction for multiple testing was done by calculating False Discovery Rate-corrected *Q*-values [32]. The significance threshold was met if the False Discovery Rate-adjusted *Q* was < 0.05.

#### Post-hoc analyses of the extreme group comparisons

Based on the reported negative genetic correlations between AN and BMI [4], we also examined whether the BMI-PGS mitigates the effect of the AN-PGS. Therefore, we carried out post-hoc analyses, comparing the above extreme groups in order to understand how they differed from one another. Post-hoc comparisons were corrected for multiple testing using Tukey’s adjustment.

## Results

### Sample Description

Following quality control of the genetic data, a total of 8,654 children with genotyping data and at least one outcome measure were included in the analyses (**Table 1**; **Table S1**; **Figure S2**).

### Association of the AN-PGS with growth trajectories (*Table 2*)

We observed several significant associations between the AN-PGS and growth trajectories (on the additive scale) exclusively in the periods of linear growth in female participants (**Table 2**). For female participants, between the ages 6.5 and 24 years, a one SD increase in the AN-PGS was associated with a 0.004% slower growth per year in their BMI trajectory given their BMI at 4 months (the intercept).

**Table 2:** Associations of the anorexia nervosa polygenic score with body composition stratified for biological sex in the Avon Longitudinal Study of Parents and Children (ALSPAC) ^a^

|  |  | Female |  |  | Male |  |  |
| --- | --- | --- | --- | --- | --- | --- | --- |
| Trajectory | Parameter | Beta <sup>b</sup> | % change <sup>c</sup> | Q <sup>d</sup> | Beta <sup>b</sup> | % change <sup>c</sup> | Q <sup>d</sup> |
| BMI <sup>e</sup> | Slope age 4 months - 1 year | 1.0000 (1.000, 1.0000) | < 0.001 | 0.76 | 1.0000 (1.0000, 1.0000) | < 0.001 | 0.71 |
| BMI | Slope age 1 year - 6.5 years | 1.0000 (1.0000, 1.0001) | < 0.001 | 0.36 | 1.0000 (1.0000, 1.0000) | < -0.001 | 0.74 |
| BMI | Slope age 6.5 years - 24 years | 1.0000 (0.9999, 1.0000) | -0.004 | **<0.001\*** | 1.0000 (1.0000, 1.0000) | < -0.001 | 0.06 |
| FMI <sup>f</sup> | Slope age 10 years - 24 years | 0.9919 (0.9774, 1.0067) | -0.81 | 0.36 | 0.9942 (0.9758, 1.0130) | -0.58 | 0.62 |
| LMI <sup>f</sup> | Slope age 10 years - 24 years | 0.9973 (0.9944, 1.0002) | -0.27 | 0.11 | 0.9993 (0.9961, 1.0027) | -0.07 | 0.74 |
| BMD <sup>f</sup> | Slope age 10 years - 24 years | 0.9960 (0.9929, 0.9991) | -0.40 | **0.02\*** | 0.9986 (0.9950, 1.0022) | -0.14 | 0.51 |
| Weight <sup>f</sup> | Slope age 10 years - 24 years | 0.9937 (0.9867, 1.0008) | -0.63 | 0.13 | 0.9956 (0.9877, 1.0036) | -0.44 | 0.36 |
| Height <sup>f</sup> | Slope age 10 years - 24 years | 0.9990 (0.9974, 1.0006) | -0.10 | 0.31 | 0.9991 (0.9970, 1.0012) | -0.09 | 0.48 |

BMI = body mass index (weight in kilograms/height<sup>2</sup> in meters); FMI = fat mass index (fat mass in kilograms/height<sup>2</sup> in meters); LMI = lean mass index (lean mass in kilograms/height<sup>2</sup> in meters); BMD = bone mineral density (gram/cm<sup>2</sup>).
<sup>a</sup> Full description of the Avon Longitudinal Study of Parents and Children is described elsewhere; [16–20].
<sup>b</sup> Betas reflect one standard deviation change in the standardized (to mean zero and standard deviation of one) BMI-PGS.
<sup>c</sup> Considering that the outcomes were log-transformed we report the percent change in the outcome for one SD increase in the PGS to ease the interpretation of the betas.
<sup>d</sup> Benjamini & Hochberg False Discovery Rate adjustment for the number tests performed [32].
<sup>e</sup> BMI trajectory was derived using spline modeling. Prior to deriving the trajectory, BMI was transformed using the natural logarithm. For the spline modeling three spline segment (Slopes) were created: a slope capturing growth between age 4 months and 1 year; a slope capturing growth between age 1 year and 6.5 years; and a slope capturing growth between age 6.5 year and 24 years.
<sup>f</sup> FMI, LMI, BMD, weight, and height trajectories were analyzed using generalized linear mixed models [33]. The anorexia nervosa PGS and the first four ancestry-informative principal components were added as fixed effects. The intercept and the slope were all allowed to vary randomly across individuals.
\* Significant after accounting for multiple testing using the false discovery rate-corrected *Q*-values. Significance was set at *Q* < 0.05.

Furthermore, in female participants, on average a one SD higher AN-PGS was associated with a 0.40 % slower growth per year for their BMD trajectory between the ages 10 and 24 years, given BMD at 10 years. There was no significant association between the AN-PGS and growth trajectories in male participants.

### Association of the BMI-PGS with growth trajectories (*Table 3*)

The BMI-PGS was associated with periods of linear growth of the BMI trajectory between the ages 1-24 years, a one SD higher BMI-PGS was associated with a 0.02% faster growth in BMI per year in female and male participants.

**Table 3:** Associations of the body mass index polygenic score with body composition stratified for biological sex in the Avon Longitudinal Study of Parents and Children ^a^

|  |  | Female |  |  | Male |  |  |
| --- | --- | --- | --- | --- | --- | --- | --- |
| Trajectory | Parameter | Beta <sup>b</sup> | % change <sup>c</sup> | Q <sup>d</sup> | Beta <sup>b</sup> | % change <sup>c</sup> | Q <sup>d</sup> |
| BMI <sup>e</sup> | Slope age 4 months - 1 year | 1.0000 (1.0000, 1.0000) | <0.001 | <0.001* | 1.0000 (1.0000, 1.0000) | <0.001 | <0.001* |
| BMI | Slope age 1 year - 6.5 years | 1.0002 (1.0001, 1.0002) | 0.02 | <0.001* | 1.0002 (1.0002, 1.0002) | 0.02 | <0.001* |
| BMI | Slope age 6.5 years - 24 years | 1.0002 (1.0002, 1.0002) | 0.02 | <0.001* | 1.0002 (1.0001, 1.0002) | 0.02 | <0.001* |
| FMI <sup>f</sup> | Slope age 10 years - 24 years | 1.1673 (1.1516, 1.1832) | 16.73 | <0.001* | 1.2040 (1.1837, 1.2247) | 20.4 | <0.001* |
| LMI <sup>f</sup> | Slope age 10 years - 24 years | 1.0211 (1.0183, 1.0239) | 2.11 | <0.001* | 1.0185 (1.0153, 1.0217) | 1.85 | <0.001* |
| BMD <sup>f</sup> | Slope age 10 years - 24 years | 1.0164 (1.0133, 1.0195) | 1.64 | <0.001* | 1.0142 (1.0107, 1.0178) | 1.42 | <0.001* |
| Weight <sup>f</sup> | Slope age 10 years - 24 years | 1.0642 (1.0572, 1.0713) | 6.42 | <0.001* | 1.0584 (1.0506, 1.0663) | 5.84 | <0.001* |
| Height <sup>f</sup> | Slope age 10 years - 24 years | 1.0016 (1.0000, 1.0033) | 0.16 | 0.09 | 1.0023 (1.0003, 1.0044) | 0.23 | 0.05 |

BMI = body mass index (weight in kilograms/height<sup>2</sup> in meters); FMI = fat mass index (fat mass in kilograms/height<sup>2</sup> in meters); LMI = lean mass index (lean mass in kilograms/height<sup>2</sup> in meters); BMD = bone mineral density (gram/cm<sup>2</sup>).
<sup>a</sup> Full description of the Avon Longitudinal Study of Parents and Children is described elsewhere; [16–20].
<sup>b</sup> Betas reflect one standard deviation change in the standardized (to mean zero and standard deviation of one) BMI-PGS.
<sup>c</sup> Considering that the outcomes were log-transformed we report the percent change in the outcome for one SD increase in the PGS to ease the interpretation of the betas.
<sup>d</sup> Benjamini & Hochberg False Discovery Rate adjustment for the number tests performed [32].
<sup>e</sup> BMI trajectory was derived using spline modeling. Prior to deriving the trajectory, BMI was transformed using the natural logarithm. For the spline modeling three spline segment (Slopes) were created: a slope capturing growth between age 4 months and 1 year; a slope capturing growth between age 1 year and 6.5 years; and a slope capturing growth between age 6.5 year and 24 years.
<sup>f</sup> FMI, LMI, BMD, weight, and height trajectories were analyzed using generalized linear mixed models [33]. The BMI-PGS and the first four ancestry-informative principal components were added as fixed effects. The intercept and the slope were all allowed to vary randomly across individuals.
\* Significant after accounting for multiple testing using the false discovery rate-corrected Q-values. Significance was set at $Q < 0.05$ .

Between the ages 10 and 24 years, a one SD higher BMI-PGS was associated with a faster growth in FMI (16.74%), LMI (2.11%), BMD (1.64%), and weight (6.42%) in female participants. We observed no association between the BMI-PGS and the height trajectory in females. We observed a similar pattern of results in male participants with a notable exception for the FMI trajectory in which a one SD higher BMI-PGS was associated with a 20.4% faster growth in FMI between the ages 10 and 24 years.

### Extreme group comparisons (*Figure 1, Table 4*)

The effects of PGS are often most discernable in the most extreme deciles so we therefore tested whether groups characterized by a particularly high PGS load differ from one another. Note that this differs from testing a formal interaction between the AN-PGS and the BMI-PGS as our focus is on understanding difference in growth at the extreme end of the PGS distribution. For the extreme group comparisons, for the BMI trajectory, we only focused on the period of growth between the ages 6.5-24 years as the AN-PGS was only associated with this stage of growth (see **Table 2**). For the FMI, LMI, BMD, weight, and height trajectories, we analyzed the period between the ages 10-24 years. Sample sizes of the extreme group comparisons can be found in **Table S2**.

**Figure 1:**
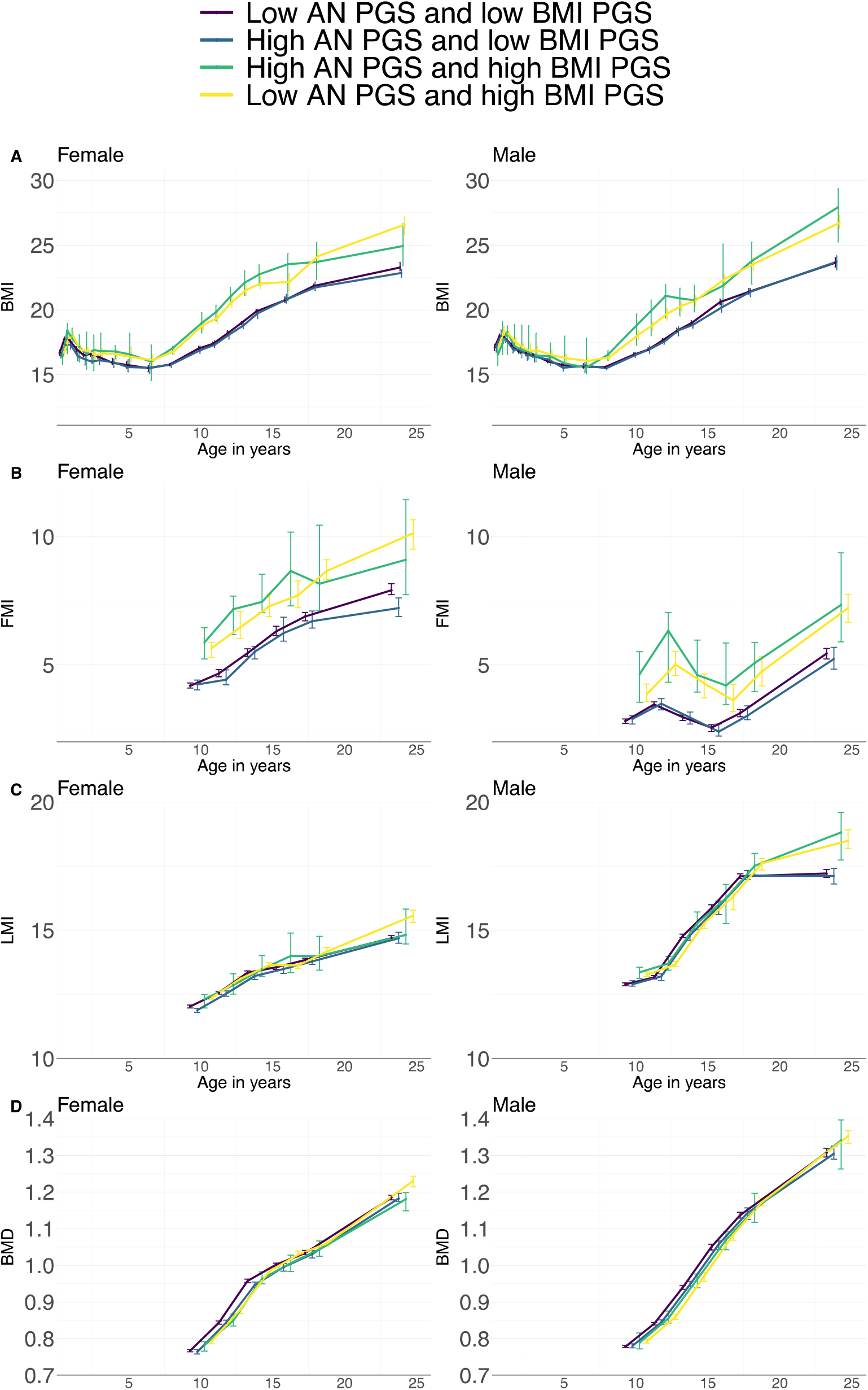
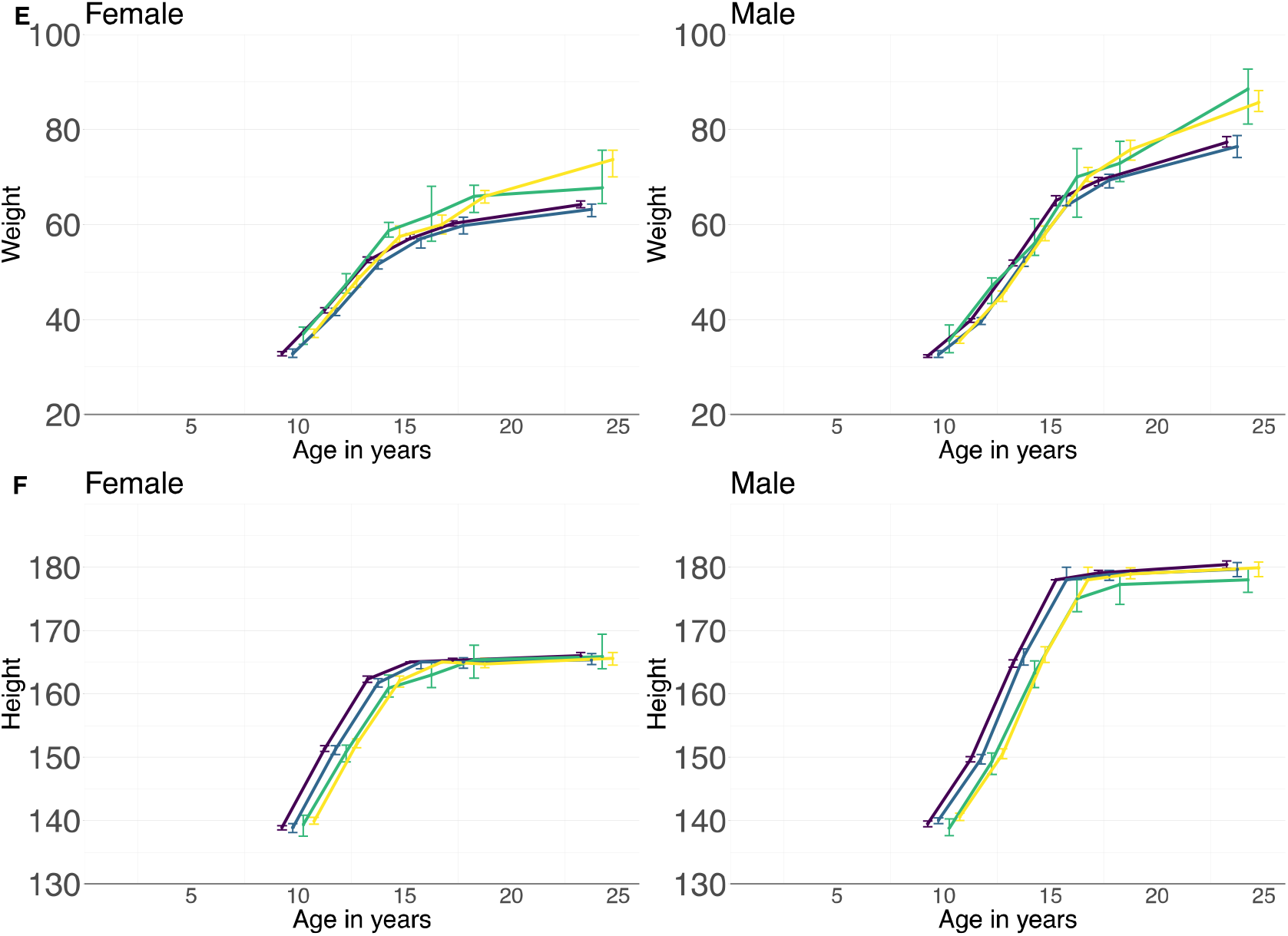
Growth trajectories for the categorial anorexia nervosa (AN) and BMI polygenic score (PGS) groups. PGS groups were derived by first dichotomizing the AN and the BMI PGS based on a cut-off value of the 8th decile of the scores. Individuals with a PGS lower than the 8th decile were grouped into a “low PGS” group, while those with a PGS score at or higher than 8th decile were grouped into a “high PGS” group. Based on this grouping we determined four categories; individuals with (1) low PGS for both AN and BMI, (2) high PGS for AN and low PGS for BMI, (3) low PGS for AN and high PGS for BMI, and (4) high PGS for both AN and BMI. **A**. Median (with 95% bootstrapped confidence interval) body mass index (BMI; weight in kilograms/height^2^ in meters) trajectories across childhood and adolescence. **B**. Median (with 95% bootstrapped confidence interval) fat mass index (FMI; fat mass in kilograms/height^2^ in meters) trajectories across childhood and adolescence. **C**. Median (with 95% bootstrapped confidence interval) lean mass index (LMI; lean mass in kilograms/height^2^ in meters) trajectories across childhood and adolescence. **D**. Median (with 95% bootstrapped confidence interval) bone mineral density (BMD) trajectories across childhood and adolescence. **E**. Median (with 95% bootstrapped confidence interval) weight (in kilograms) trajectories across childhood and adolescence. **F**. Median (with 95% bootstrapped confidence interval) height (in centimeters) trajectories across childhood and adolescence.

**Table 4:** Associations of the combined anorexia nervosa and body mass index polygenic score with the body composition measures stratified for biological sex using linear mixed models in the Avon Longitudinal Study of Parents and Children ^a^

|  |  | Female |  |  | Male |  |  |
| --- | --- | --- | --- | --- | --- | --- | --- |
| Trajectory | PGS category <sup>b</sup> | Beta <sup>c</sup> | % change <sup>d</sup> | Q <sup>e</sup> | Beta <sup>c</sup> | % change <sup>d</sup> | Q <sup>e</sup> |
| BMI (age 6.5 years – 24 years) | Low AN and high BMI | 1.10 (1.08, 1.11) | 9.78 | <0.001* | 1.07 (1.06, 1.09) | 7.41 | <0.001* |
| BMI (age 6.5 years – 24 years) | High AN and high BMI | 1.11 (1.08, 1.14) | 10.74 | <0.001* | 1.10 (1.08, 1.13) | 10.28 | <0.001* |
| BMI (age 6.5 years – 24 years) | High AN and low BMI | 1.10 (0.99, 1.01) | -0.17 | 0.89 | 1.00 (0.99, 1.01) | 0.09 | 0.93 |
| FMI (age 10 years – 24 years) | Low AN and high BMI | 1.28 (1.23, 1.33) | 27.81 | <0.001* | 1.38 (1.32, 1.45) | 38.14 | <0.001* |
| FMI (age 10 years – 24 years) | High AN and high BMI | 1.32 (1.21, 1.43) | 31.56 | <0.001* | 1.48 (1.33, 1.64) | 47.76 | <0.001* |
| FMI (age 10 years – 24 years) | High AN and low BMI | 0.99 (0.95, 1.02) | -1.29 | 0.65 | 1.02 (0.97, 1.07) | 1.51 | 0.71 |
| LMI (age 10 years – 24 years) | Low AN and high BMI | 1.03 (1.03, 1.04) | 3.32 | <0.001* | 1.03 (1.02, 1.03) | 2.77 | <0.001* |
| LMI (age 10 years – 24 years) | High AN and high BMI | 1.03 (1.01, 1.04) | 2.77 | 0.001* | 1.03 (1.02, 1.05) | 3.26 | <0.001* |
| LMI (age 10 years – 24 years) | High AN and low BMI | 0.99 (0.99, 1.00) | -0.56 | 0.17 | 1.00 (0.99, 1.01) | 0.00 | 0.99 |
| BMD (age 10 years – 24 years) | Low AN and high BMI | 1.03 (1.02, 1.04) | 2.92 | <0.001* | 1.03 (1.02, 1.03) | 2.77 | <0.001* |
| BMD (age 10 years – 24 years) | High AN and high BMI | 1.02 (1.01, 1.04) | 2.09 | 0.012* | 1.03 (1.02, 1.05) | 3.26 | <0.001* |
| BMD (age 10 years – 24 years) | High AN and low BMI | 0.99 (0.99, 1.00) | -0.68 | 0.07 | 1.00 (0.99, 1.01) | 0.00 | 0.99 |
| Weight (age 10 years – 24 years) | Low AN and high BMI | 1.11 (1.09, 1.13) | 11.29 | <0.001* | 1.12 (1.10, 1.13) | 11.52 | <0.001* |
| Weight (age 10 years – 24 years) | High AN and high BMI | 1.10 (1.06, 1.14) | 10.08 | <0.001* | 1.12 (1.08, 1.16) | 11.85 | <0.001* |
| Weight (age 10 years – 24 years) | High AN and low BMI | 0.99 (0.98, 1.01) | -0.77 | 0.53 | 1.00 (0.98, 1.02) | 0.17 | 0.92* |
| Height (age 10 years – 24 years) | Low AN and high BMI | 1.00 (1.00, 1.01) | 0.15 | 0.64 | 1.01 (1.00, 1.01) | 0.62 | 0.01* |
| Height (age 10 years – 24 years) | High AN and high BMI | 1.00 (0.99, 1.01) | -0.25 | 0.72 | 1.00 (0.99, 1.01) | -0.25 | 0.72 |
| Height (age 10 years – 24 years) | High AN and low BMI | 1.00 (1.00, 1.00) | -0.08 | 0.79 | 1.00 (1.00, 1.01) | 0.13 | 0.71 |

AN = anorexia nervosa; PGS = polygenic score; BMI = body mass index (weight in kilograms/height<sup>2</sup> in meters); FMI = fat mass index (fat mass in kilogram /height<sup>2</sup> in meters); LMI = lean mass index (lean mass in kilograms/height<sup>2</sup> in meters); BMD = bone mineral density (gram/cm<sup>2</sup>).
<sup>a</sup> Full description of the Avon Longitudinal Study of Parents and Children is described elsewhere; [16–20].
<sup>b</sup> Categorical variable derived from dichotomizing the AN-PGS and the BMI-PGS. For both the AN and the BMI PGS, individuals with scores at or greater than the 8<sup>th</sup> decile point were regarded as the “high PGS group” while those with scores lower were considered the “low PGS group”. From the dichotomized AN-PGS and BMI-PGS we were able to create a categorical variable with four levels (1) low AN-PGS and low BMI-PGS (2) high AN-PGS and low BMI-PGS (3) high AN-PGS and high BMI-PGS (4) low AN-PGS and high BMI-PGS. The “low AN-PGS and low BMI-PGS” group was used as the reference category in the analyses.
<sup>c</sup> Betas reflect change in outcome compared to the reference category “low AN-PGS and low BMI-PGS”.
<sup>d</sup> Considering that the outcomes were log-transformed we report the percent change in the outcome in the comparison to the reference group (“low AN-PGS and BMI-PGS”) to ease the interpretation of the betas.
<sup>e</sup> Benjamini & Hochberg False Discovery Rate adjusting for the number of hypothesis tested [32].
\* Significant after accounting for multiple testing using the False Discovery Rate-corrected Q-values. Significance was set at $Q < 0.05$ .

Individuals with a low AN-PGS and a high BMI-PGS followed on average faster growth than individuals with a low AN-PGS and a low BMI-PGS. The difference was most pronounced for the FMI trajectory (**Table 4**). Female participants with a low AN-PGS and a high BMI-PGS had a 27.81% faster growth in FMI compared to their peers with a low AN-PGS and a low BMI-PGS. Male participants with a low AN-PGS and a high BMI-PGS also showed faster growth in FMI (38.14%) compared to their peers with a low AN-PGS and a low BMI-PGS. Furthermore, although not statistically significant, we observed a trend in which female participants with high AN-PGS and a low BMI-PGS had slower growth for the BMI, FMI, LMI, BMD, weight, and height trajectories compared to the reference category of individuals with low AN-PGS and low BMI-PGS (**Figure 1, Table 4**).

### Post-hoc analyses extreme group comparisons (*Table 5*)

In order to understand how the different PGS groups differ from one another, we conducted post-hoc analyses of the extreme group comparisons. Though the high AN-PGS/low BMI-PGS did not differ from the reference category (**Table 4**), this group did differ (slower growth) significantly from the high AN-PGS/high BMI-PGS group and the low AN-PGS/high BMI-PGS group (**Table 5**; **Table S3**).

**Table 5:**
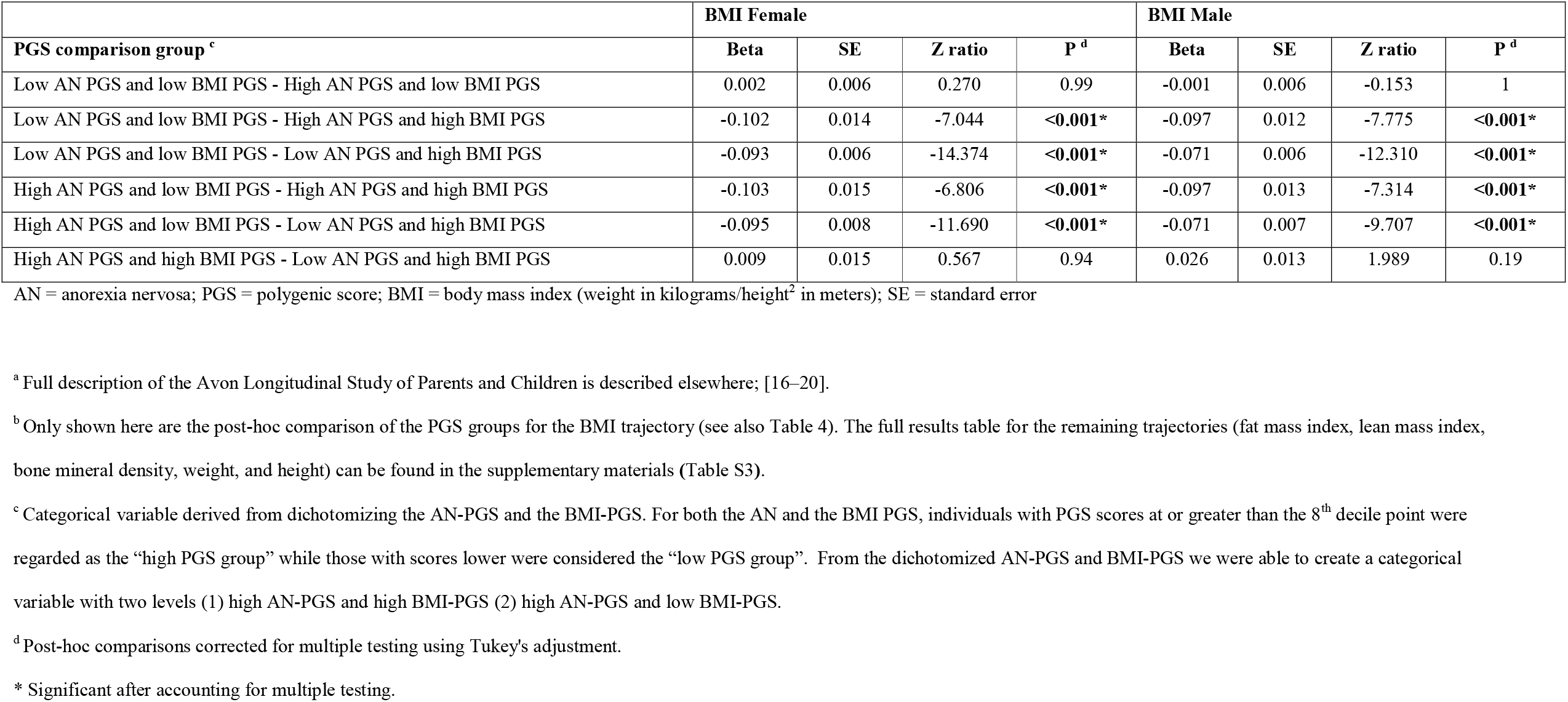
Associations of the combined anorexia nervosa and body mass (BMI) index polygenic score with the body composition measures stratified for biological sex using linear mixed models in the Avon Longitudinal Study of Parents and Children: Post-hoc analyses of the BMI trajectory ^a b^

## Discussion

Using a longitudinal data across the firs two decades of life, we report that common genomic variants associated with AN and BMI are significantly associated with growth trajectories. Female participants with a high AN genetic liability differ significantly in growth as marked by slower growth as early as age 6.5 years for the BMI trajectory and age 10 years for the BMD trajectory. This effect was not observed in male participants.

Sex differences in body composition have been well-documented in the medical literature with men on average having more lean body mass, higher BMD, being taller, and having a lower fat mass than women [34]. These biological differences are driven by both environmental and genetic factors [31,35]. In a recent study, we reported a negative genetic correlation between body fat percentage and AN, which was significantly more pronounced in women than in men (women SNP-*r*_g_ = −0.44, SE = 0.04; men SNP-*r*_g_ = −0.26; SE = 0.04) [35]. The negative association between the AN-PGS and the BMD trajectory is consistent with the established literature that AN is phenotypically associated with BMD; AN has marked and severe adverse effects on bone metabolism [3,36]. The negative association between the AN-PGS and BMD was only found in female participants. The observed sex-specific effects of the AN-PGS suggest that a specific set of common genetic variants may be differentially active in women and may increase the liability for AN. These results also underscore the importance of collecting adequate samples from men with AN and all EDs, allowing for the calculating ED PGS specific for men, to ensure our ability to identify differential genetic effects.

We also demonstrate that developmental changes in body composition are in part driven by the BMI-PGS as early as 4 months of age. The pattern of association of the BMI-PGS with the growth trajectories did not differ between the sexes. The BMI-PGS was significantly associated with FMI and LMI trajectories whereas the AN-PGS was not— another point of divergence between the BMI-PGS and the AN-PGS. The association of the BMI-PGS with FMI and LMI is in part due to shared genomics [35]. In a previous study [35], we showed a significant genetic correlation between childhood BMI and adiposity (SNP-*r*_g_ = 0.46) and between childhood BMI and lean mass (SNP-*r*_g_ = 0.41). Consistent with our findings, increases in body weight are reported to be associated with higher BMD [37]. Whether these associations are due to adaptive changes of the body to increased body weight (i.e., higher BMI-PGS leads to increases in body weight, which in turn leads to increases in BMD to sustain a higher body weight) or whether this is due to shared genomics is currently unclear.

We also sought to understand how AN and BMI genetics could co-influence growth across childhood and adolescence. We found that female participants with high AN-PGS and low BMI-PGS did not significantly differ from a reference group with a low AN-PGS and low BMI-PGS, although the direction of difference aligned with expectations—individuals with high AN-PGS and low BMI-PGS have lower growth trajectories compared to the reference group. Interestingly, female participants with high genetic liability for AN and low genetic liability for BMI followed lower growth trajectories than individuals with high PGS on both traits, which might suggest that the BMI-PGS mitigated the effects of the AN-PGS. This interpretation is consistent with reports of a negative genetic correlation between AN and BMI [4,38]. Taken together, the findings from the univariate AN-PGS analyses and the findings from the extreme-group comparisons suggest that genetic liability to high weight exerts a modulating role in the association between the AN-PGS and growth trajectories. However, we cannot exclude that other PGS (e.g., depression) could also exert influence on these associations [39].

The observation that AN-PGS and BMI-PGS co-influence growth trajectories is a novel finding that encourages exploration of underlying biological mechanisms. The fact that a PGS of one trait could mitigate the effects of another trait is intriguing and has previously been described in a study investigating stressful life events; higher well-being PGS buffered against increased depressive symptoms following a spouse’s death [40]. Common genomic variants that are associated with AN or BMI are primarily expressed in the central nervous system [4,13,41], suggesting that body mass is behaviorally influenced. This view is also supported by our previous work and that of others, showing that a PGS for a higher BMI is associated with several adolescent eating behaviors including higher propensity to engage in self-reported binge eating [42–44]. Taken together, several sources of converging evidence suggest that polygenic risk for AN and/or BMI may impact growth at least in part via eating behaviors.

This study has several strengths including the large sample size and the prospective and repeated collection of objective body composition measures spanning more than 20 years. We included a negative control (i.e., the height trajectory, which was not associated with the AN-PGS), adequately controlled for multiple testing, and controlled for potential genetic confounders using ancestry informative principal components. The use of splines enabled modeling of BMI trajectories more accurately as growth throughout childhood is not linear. Further, by stratifying on biological sex, we were able to identify sex-related effects that otherwise would have been masked, and censoring on eating disorders ruled out that growth changes during puberty were a consequence of an eating disorder.

Findings from this study should be interpreted in the context of some limitations. Participants were recruited from the same geographical region in the south-west of England and, therefore, the results may not be generalizable to other populations. However, the homogeneity of this sample lends itself to genetic analyses as bias from population stratification is low [45]. Considering the longitudinal nature of the study, participants tend to drop out over time leading to missing data. We maximized available data by using mixed effects models, which allowed us to use all available data points in deriving the growth trajectories rather than only including complete cases. We acknowledge that any bias as a consequence of missing data in our analyses could have biased our results towards the null. The effect sizes observed in this study were relatively small, but are consistent with those previously reported in other PGS studies and aid our understanding of growth trajectories [15,46]. Height and weight data in ALSPAC were obtained from a range of clinical sources, which could have introduced variability in the obtained measures. However, measures of height and weight were highly correlated across different measurement settings (clinic and self-report) (**Figure S1**). Furthermore, the BMI GWAS that was used in deriving the PGS was conducted mostly in adult participants, which could have biased our analyses. However, for BMI, a previous study found substantial overlap between childhood and adult BMI GWAS loci (R_g_ = 0.76, P = 1.45 × 10^−112^) [47]. Regarding the AN GWAS, we would like to also emphasize that both adolescents and children were included in the study; therefore, the AN GWAS should be equipped to pick up genomic variants associated with the disorder in adolescence which is the typical age of onset.

In conclusion, we show that polygenic risk for AN and BMI have detectable sex-specific effects on growth during the first two decades of life. Especially, female participants with high polygenic risk for AN and a low polygenic risk for BMI likely constitute a high-risk group as they followed lower growth trajectories, which have previously been associated with AN in the ALSPAC sample [7]. This study adds to a growing body of evidence suggesting that risk for AN could emerge during early childhood and that a combination of AN and BMI polygenic risk could aid the early identification of individuals at high risk for AN. These findings encourage further research in understanding how the AN-PGS and the BMI-PGS co-influence growth during childhood in which the BMI-PGS can amplify or mitigate the effects of the AN-PGS.

## Data Availability

Please note that the study website contains details of all the data that is available through a fully searchable data dictionary and variable search tool" and reference the following webpage: http://www.bristol.ac.uk/alspac/researchers/our-data/.

## Data Availability

All data produced in the present study are available upon reasonable request to the authors

## Acknowledgements

We are extremely grateful to all the families who took part in this study, the midwives for their help in recruiting them, and the whole ALSPAC team, which includes interviewers, computer and laboratory technicians, clerical workers, research scientists, volunteers, managers, receptionists and nurses.

## Funding

This study represents independent research part funded by the UK National Institute for Health Research (NIHR) Biomedical Research Centre at South London and Maudsley NHS Foundation Trust and King’s College London. The views expressed are those of the author(s) and not necessarily those of the UK NHS, the NIHR or the Department of Health. High performance computing facilities were funded with capital equipment grants from the GSTT Charity (TR130505) and Maudsley Charity (980). This work was supported by the UK Medical Research Council and the Medical Research Foundation (ref: MR/R004803/1). The UK Medical Research Council and Wellcome (Grant ref: 102215/2/13/2 and 217065/Z/19/Z) and the University of Bristol provide core support for ALSPAC. A comprehensive list of grants funding is available on the ALSPAC website (http://www.bristol.ac.uk/alspac/external/documents/grant-acknowledgements.pdf); This research was specifically funded by the NIHR (CS/01/2008/014), the NIH (MH087786-01). GWAS data was generated by Sample Logistics and Genotyping Facilities at Wellcome Sanger Institute and LabCorp (Laboratory Corporation of America) using support from 23andMe. NM and CB acknowledge funding from the National Institute of Mental Health (R21 MH115397). CMB is supported by NIMH (R01MH120170; R01MH124871; R01MH119084; R01MH118278; R01 MH124871); Brain and Behavior Research Foundation Distinguished Investigator Grant; Swedish Research Council (Vetenskapsrådet, award: 538-2013-8864); Lundbeck Foundation (Grant no. R276-2018-4581). The content is solely the responsibility of the authors and does not necessarily represent the official views of the National Institutes of Health. The funders were not involved in the design or conduct of the study; collection, management, analysis, or interpretation of the data; or preparation, review, or approval of the manuscript

## Conflict of interest

Dr. Breen has received grant funding from and served as a consultant to Eli Lilly, has received honoraria from Illumina and has served on advisory boards for Otsuka. Dr. Bulik is a grant recipient from and has served on advisory boards for Shire and is a consultant for Idorsia. She receives royalties from Pearson. She is on the Clinical Advisory Board of Equip Health Inc. All other authors have indicated they have no conflicts of interest to disclose.

## Author Contribution

MA, CH, and MH analyzed the data. MA, CH, and MH drafted the manuscript. NM, CMB, RFL, and GB supervised the work. All authors substantially contributed to the conception and interpretation of the work, revised the manuscript for important intellectual content and approved the final version. All authors agree to be accountable for all aspects of this work.

## Notes

### Clinical Trial

ALSPAC is a longitudinal birth cohort and as such is not a clinical trial. The study has been approved by the Research Ethics Committee. The initial approvals of the study were:
1. Bristol and Weston Health Authority: E1808 Children of the Nineties: Avon
Longitudinal Study of Pregnancy and Childhood (ALSPAC). (28th November
1989)
2. Southmead Health Authority: 49/89 Children of the Nineties -
"ALSPAC". (5th April 1990)
3. Frenchay Health Authority: 90/8 Children of the Nineties. (28th June 1990)
For more details regarding ethical approval please visit: http://www.bristol.ac.uk/alspac/researchers/research-ethics/.

### Author Declarations

Ethical approval for the study was obtained from the ALSPAC Ethics and Law Committee and the Local Research Ethics Committees (Bristol and Weston Health Authority: E1808 Children of the Nineties: ALSPAC, 28th November 1989 (for details see: www.bristol.ac.uk/alspac/researchers/research-ethics/).

### Summary of Updates

In this version of the manuscript, we use the more powerful BMI polygenic scores rather than the obesity PGS

## References

1. Treasure J, Zipfel S, Micali N, Wade T, Stice E, Claudino A, et al. Anorexia nervosa. Nat Rev Dis Prim [Internet]. Macmillan Publishers Limited; 2015;1:1–22. Available from: http://dx.doi.org/10.1038/nrdp.2015.74

2. Polito A, Cuzzolaro M, Raguzzini A, Censi L, Ferro-Luzzi A. Body composition changes in anorexia nervosa. Eur J Clin Nutr [Internet]. 1998;52:655–62. Available from: http://www.nature.com/articles/1600618

3. Hübel C, Yilmaz Z, Schaumberg KE, Breithaupt L, Hunjan A, Horne E, et al. Body composition in anorexia nervosa: Meta-analysis and meta-regression of cross-sectional and longitudinal studies. Int J Eat Disord [Internet]. 2019;52:1205–23. Available from: https://onlinelibrary.wiley.com/doi/abs/10.1002/eat.23158

4. Watson HJ, Yilmaz Z, Thornton LM, Hübel C, Coleman JRII, Gaspar HA, et al. Genome-wide association study identifies eight risk loci and implicates metabo-psychiatric origins for anorexia nervosa. Nat Genet [Internet]. 2019;51:1207–14. Available from: http://www.nature.com/articles/s41588-019-0439-2

5. Stice E. Interactive and Mediational Etiologic Models of Eating Disorder Onset: Evidence from Prospective Studies. Annu Rev Clin Psychol. 2016;

6. Tyrka AR, Waldron I, Graber JA, Brooks-Gunn J. Prospective predictors of the onset of anorexic and bulimic syndromes. Int J Eat Disord. 2002;

7. Yilmaz Z, Gottfredson NC, Zerwas SC, Bulik CM, Micali N. Developmental Premorbid Body Mass Index Trajectories of Adolescents With Eating Disorders in a Longitudinal Population Cohort. J Am Acad Child Adolesc Psychiatry [Internet]. Elsevier; 2019;58:191–9. Available from: http://dx.doi.org/10.1016/j.jaac.2018.11.008

8. Simon GE, Von Korff M, Saunders K, Miglioretti DL, Crane PK, Van Belle G, et al. Association between obesity and psychiatric disorders in the US adult population. Arch Gen Psychiatry. 2006;63:824–30.

9. González-Muniesa P, Mártinez-González M-A, Hu FB, Després J-P, Matsuzawa Y, Loos RJF, et al. Obesity. Nat Rev Dis Prim [Internet]. 2017;3:17034. Available from: http://www.nature.com/articles/nrdp201734

10. Tomiyama AJ, Carr D, Granberg EM, Major B, Robinson E, Sutin AR, et al. How and why weight stigma drives the obesity “epidemic” and harms health. BMC Med. 2018;

11. Spahlholz J, Baer N, König HH, Riedel-Heller SG, Luck-Sikorski C. Obesity and discrimination - a systematic review and meta-analysis of observational studies. Obes. Rev. 2016.

12. Daly M, Sutin AR, Robinson E. Perceived Weight Discrimination Mediates the Prospective Association Between Obesity and Physiological Dysregulation: Evidence From a Population-Based Cohort. Psychol Sci. 2019;

13. Locke AE, Kahali B, Berndt SI, Justice AE, Pers TH, Day FR, et al. Genetic studies of body mass index yield new insights for obesity biology. Nature. 2015;518:197–206.

14. Yengo L, Sidorenko J, Kemper KE, Zheng Z, Wood AR, Weedon MN, et al. Meta-analysis of genome-wide association studies for height and body mass index in ∼700000 individuals of European ancestry. Hum Mol Genet [Internet]. 2018;27:3641–9. Available from: https://academic.oup.com/hmg/article/27/20/3641/5067845

15. Khera A V., Chaffin M, Wade KH, Zahid S, Brancale J, Xia R, et al. Polygenic Prediction of Weight and Obesity Trajectories from Birth to Adulthood. Cell [Internet]. Elsevier Inc.; 2019;177:587–596.e9. Available from: https://doi.org/10.1016/j.cell.2019.03.028

16. Fraser A, Macdonald-wallis C, Tilling K, Boyd A, Golding J, Davey smith G, et al. Cohort profile: The avon longitudinal study of parents and children: ALSPAC mothers cohort. Int J Epidemiol. 2013;42:97–110.

17. Boyd A, Golding J, Macleod J, Lawlor DA, Fraser A, Henderson J, et al. Cohort profile: The ‘Children of the 90s’-The index offspring of the avon longitudinal study of parents and children. Int J Epidemiol. 2013;42:111–27.

18. Golding, Pembrey, Jones, The Alspac Study Team. ALSPAC-The Avon Longitudinal Study of Parents and Children. Paediatr Perinat Epidemiol [Internet]. 2001;15:74–87. Available from: http://doi.wiley.com/10.1046/j.1365-3016.2001.00325.x

19. Golding J. The Avon Longitudinal Study of Parents and Children (ALSPAC)--study design and collaborative opportunities. Eur J Endocrinol [Internet]. 2004;U119–23. Available from: https://eje.bioscientifica.com/view/journals/eje/151/Suppl_3/U119.xml

20. Northstone K, Lewcock M, Groom A, Boyd A, Macleod J, Timpson N, et al. The Avon Longitudinal Study of Parents and Children (ALSPAC): an update on the enrolled sample of index children in 2019. Wellcome Open Res [Internet]. 2019;4:51. Available from: https://wellcomeopenresearch.org/articles/4-51/v1

21. Harris PA, Taylor R, Thielke R, Payne J, Gonzalez N, Conde JG. Research electronic data capture (REDCap)-A metadata-driven methodology and workflow process for providing translational research informatics support. J Biomed Inform. 2009;

22. Harris PA, Taylor R, Minor BL, Elliott V, Fernandez M, O’Neal L, et al. The REDCap consortium: Building an international community of software platform partners. J. Biomed. Inform. 2019.

23. Micali N, De Stavola B, Ploubidis G, Simonoff E, Treasure J, Field AE. Adolescent eating disorder behaviours and cognitions: Gender-specific effects of child, maternal and family risk factors. Br J Psychiatry [Internet]. 2015;207:320–7. Available from: https://www.cambridge.org/core/product/identifier/S0007125000239469/type/journal_article

24. Micali N, Solmi F, Horton NJ, Crosby RD, Eddy KT, Calzo JP, et al. Adolescent Eating Disorders Predict Psychiatric, High-Risk Behaviors and Weight Outcomes in Young Adulthood. J Am Acad Child Adolesc Psychiatry [Internet]. Elsevier Inc; 2015;54:652–659.e1. Available from: http://dx.doi.org/10.1016/j.jaac.2015.05.009

25. Hübel C, Abdulkadir M, Herle M, Loos RJF, Breen G, Bulik CM, et al. One size does not fit all. Genomics differentiates among anorexia nervosa, bulimia nervosa, and binge-eating disorder. Int J Eat Disord [Internet]. 2021;54:785–93. Available from: https://onlinelibrary.wiley.com/doi/10.1002/eat.23481

26. Warrington NM, Howe LD, Wu YY, Timpson NJ, Tilling K, Pennell CE, et al. Association of a Body Mass Index Genetic Risk Score with Growth throughout Childhood and Adolescence. Sleegers K, editor. PLoS One [Internet]. 2013;8:e79547. Available from: https://dx.plos.org/10.1371/journal.pone.0079547

27. Ge T, Chen CY, Ni Y, Feng YCA, Smoller JW. Polygenic prediction via Bayesian regression and continuous shrinkage priors. Nat Commun [Internet]. Springer US; 2019;10:1–10. Available from: http://dx.doi.org/10.1038/s41467-019-09718-5

28. Kahn RS, Sommer IE, Murray RM, Meyer-Lindenberg A, Weinberger DR, Cannon TD, et al. Schizophrenia. Nat Rev Dis Prim [Internet]. 2015;1:15067. Available from: http://www.nature.com/articles/nrdp201567

29. Bates D, Mächler M, Bolker BM, Walker SC. Fitting Linear Mixed-Effects Models Using lme4. 2015;67.

30. Hu□bel C, Gaspar HA, Coleman JRI, Hanscombe KB, Purves K, Prokopenko I, et al. Genetic correlations of psychiatric traits with body composition and glycemic traits are sex-and age-dependent. Nat Commun [Internet]. 2019;10:5765. Available from: http://www.nature.com/articles/s41467-019-13544-0

31. Zillikens MC, Yazdanpanah M, Pardo LM, Rivadeneira F, Aulchenko YS, Oostra BA, et al. Sex-specific genetic effects influence variation in body composition. Diabetologia. 2008;51:2233–41.

32. Benjamini Y, Hochberg Y. Controlling the False Discovery Rate□: a Practical and Powerful Approach to Multiple Testing. J R Stat Soc Ser B. 1995;57:289–300.

33. Herle M, Micali N, Abdulkadir M, Loos R, Bryant-Waugh R, Hübel C, et al. Identifying typical trajectories in longitudinal data: modelling strategies and interpretations. Eur J Epidemiol [Internet]. Springer Netherlands; 2020;35:205–22. Available from: https://doi.org/10.1007/s10654-020-00615-6

34. Wells JCK. Sexual dimorphism of body composition. Best Pract. Res. Clin. Endocrinol. Metab. 2007.

35. Hübel C, Gaspar HA, Coleman JRII, Finucane H, Purves KL, Hanscombe KB, et al. Genomics of body fat percentage may contribute to sex bias in anorexia nervosa. Am J Med Genet Part B Neuropsychiatr Genet [Internet]. 2019;180:428–38. Available from: http://doi.wiley.com/10.1002/ajmg.b.32709

36. Bulik CM, Carroll IM, Mehler P. Endocrinology & Metabolism Reframing anorexia nervosa as a metabo-psychiatric disorder. Trends Endocrinol Metab [Internet]. The Authors; 2021;xx:1–10. Available from: https://doi.org/10.1016/j.tem.2021.07.010

37. Maïmoun L, Garnero P, Mura T, Nocca D, Lefebvre P, Philibert P, et al. Specific Effects of Anorexia Nervosa and Obesity on Bone Mineral Density and Bone Turnover in Young Women. J Clin Endocrinol Metab [Internet]. 2020;105:e1536–48. Available from: https://academic.oup.com/jcem/advance-article/doi/10.1210/clinem/dgz259/5672712

38. Duncan L, Yilmaz Z, Gaspar H, Walters R, Goldstein J, Anttila V, et al. Significant Locus and Metabolic Genetic Correlations Revealed in Genome-Wide Association Study of Anorexia Nervosa. Am J Psychiatry [Internet]. 2017;174:850–8. Available from: http://ajp.psychiatryonline.org/doi/10.1176/appi.ajp.2017.16121402

39. Pine DS, Goldstein RB, Wolk S, Weissman MM. The Association Between Childhood Depression and Adulthood Body Mass Index. Pediatrics [Internet]. 2001;107:1049–56. Available from: http://pediatrics.aappublications.org/cgi/doi/10.1542/peds.107.5.1049

40. Domingue BW, Liu H, Okbay A, Belsky DW. Genetic Heterogeneity in Depressive Symptoms Following the Death of a Spouse: Polygenic Score Analysis of the U.S. Health and Retirement Study. Am J Psychiatry [Internet]. 2017;174:963–70. Available from: http://ajp.psychiatryonline.org/doi/10.1176/appi.ajp.2017.16111209

41. Goodarzi MO. Genetics of obesity: what genetic association studies have taught us about the biology of obesity and its complications. Lancet Diabetes Endocrinol [Internet]. Elsevier Ltd; 2018;6:223–36. Available from: http://dx.doi.org/10.1016/S2213-8587(17)30200-0

42. Abdulkadir M, Herle M, De Stavola BL, Hübel C, Santos Ferreira DL, Loos RJF, et al. Polygenic Score for Body Mass Index Is Associated with Disordered Eating in a General Population Cohort. J Clin Med [Internet]. 2020;9:1187. Available from: https://www.mdpi.com/2077-0383/9/4/1187

43. Nagata JM, Braudt DB, Domingue BW, Bibbins-Domingo K, Garber AK, Griffiths S, et al. Genetic risk, body mass index, and weight control behaviors: Unlocking the triad. Int J Eat Disord [Internet]. 2019;52:825–33. Available from: https://onlinelibrary.wiley.com/doi/abs/10.1002/eat.23083

44. Robinson L, Zhang Z, Jia T, Bobou M, Roach A, Campbell I, et al. Association of Genetic and Phenotypic Assessments With Onset of Disordered Eating Behaviors and Comorbid Mental Health Problems Among Adolescents. JAMA Netw open. 2020;3:e2026874.

45. Hellwege JN, Keaton JM, Giri A, Gao X, Velez Edwards DR, Edwards TL. Population Stratification in Genetic Association Studies. Curr Protoc Hum Genet. 2017;95:1.22.1-1.22.23.

46. Stergiakouli E, Martin J, Hamshere ML, Heron J, St Pourcain B, Timpson NJ, et al. Association between polygenic risk scores for attention-deficit hyperactivity disorder and educational and cognitive outcomes in the general population. Int J Epidemiol [Internet]. 2016;09:dyw216. Available from: http://ije.oxfordjournals.org/lookup/doi/10.1093/ije/dyw216

47. Vogelezang S, Bradfield JP, Ahluwalia TS, Curtin JA, Lakka TA, Grarup N, et al. Novel loci for childhood body mass index and shared heritability with adult cardiometabolic traits. Copenhaver GP, editor. PLOS Genet [Internet]. 2020;16:e1008718. Available from: https://dx.plos.org/10.1371/journal.pgen.1008718

