## Supplementary material for "The impact of anorexia nervosa and BMI polygenic risk on childhood growth: a 20-year longitudinal population-based study"

**Supplemental information**

**Genotyping quality control checks**

Individuals with disproportionate levels of individual missingness (i.e., >3%), insufficient sample replication (identity by descent < 0.1), biological sex mismatch, and non-European ancestry (as defined by multi-dimensional scaling using the HapMap Phase II, release 22, reference populations) were excluded. Single nucleotide polymorphisms (SNPs) with a minor allele frequency (MAF) of < 1%, excessive missingness (i.e., call rate < 95%), or a departure from the Hardy–Weinberg equilibrium (*P* value < 5 x 10^-7^) were removed. Imputation was conducted with Impute3 using the HRC 1.0 as the reference panel [1] and phasing was carried out using ShapeIT (v2.r644). Finally, post-imputation quality control checks were performed; any SNPs with MAF less than 1%, Impute3 information quality metric of < 0.8, and not confirming to Hardy-Weinberg equilibrium (*P* < 5 × 10^-7^) were removed. After data cleaning, a total of 8,654 individuals (4,225 females and 4,429 males) and 4,054,653 SNPs remained eligible for analyses.

**Table S1:** Descriptive of the body composition measures of the Avon Longitudinal Study of Parents and Children (ALSPAC)^a^

|  |  | Female | | Male | |
| --- | --- | --- | --- | --- | --- |
| Body composition measure | **Age (months)** | **N** | **Median (IQR)** | **N** | **Median (IQR)** |
| BMI ^b^ | 4 | 337 | 16.61 (15.76, 17.41) | 363 | 17.12 (16.02, 18.02) |
|  | 8 | 438 | 17.67 (16.76, 18.6) | 491 | 18.04 (17.01, 19.14) |
|  | 12 | 423 | 17.67 (16.69, 18.52) | 467 | 17.94 (17.05, 18.89) |
|  | 18 | 405 | 16.87 (16.03, 17.73) | 440 | 17.25 (16.38, 18.15) |
|  | 24 | 370 | 16.52 (15.75, 17.47) | 411 | 16.89 (16.06, 17.77) |
|  | 30 | 384 | 16.53 (15.70, 17.37) | 422 | 16.67 (15.89, 17.48) |
|  | 36 | 385 | 16.38 (15.53, 17.25) | 417 | 16.53 (15.77, 17.26) |
|  | 48 | 372 | 16.08 (15.23, 17.06) | 419 | 16.15 (15.52, 16.9) |
|  | 60 | 367 | 15.88 (15.03, 16.87) | 403 | 15.82 (15.17, 16.67) |
|  | 78 | 544 | 15.57 (14.49, 16.91) | 534 | 15.73 (14.86, 16.93) |
|  | 96 | 3,130 | 15.92 (14.92, 17.32) | 3,224 | 15.72 (14.88, 16.82) |
|  | 120 | 3,072 | 17.31 (15.77, 19.41) | 3,026 | 16.77 (15.61, 18.73) |
|  | 132 | 2,977 | 17.7 (16.07, 19.94) | 2,923 | 17.26 (15.93, 19.50) |
|  | 144 | 2,902 | 18.6 (16.74, 21.14) | 2,797 | 17.96 (16.43, 20.50) |
|  | 156 | 2,742 | 19.34 (17.52, 21.78) | 2,630 | 18.76 (17.04, 21.12) |
|  | 168 | 2,433 | 20.17 (18.43, 22.58) | 2,394 | 19.28 (17.71, 21.45) |
|  | 192 | 1,496 | 20.95 (19.47, 23.14) | 1,289 | 20.83 (19.14, 22.78) |
|  | 216 | 1,986 | 22.14 (20.35, 24.78) | 1,681 | 21.76 (20.08, 24.34) |
|  | 288 | 1,671 | 23.63 (21.47, 27.04) | 1,131 | 24.25 (21.97, 27.09) |
| FMI ^c^ | 120 | 2,932 | 4.40 (3.14, 6.18) | 2,880 | 2.96 (2.08, 4.67) |
|  | 144 | 2,862 | 4.91 (3.52, 7.11) | 2,750 | 3.65 (2.53, 5.90) |
|  | 168 | 2,406 | 5.77 (4.28, 7.74) | 2,353 | 3.11 (2.09, 5.26) |
|  | 192 | 1,266 | 6.50 (5.00, 8.36) | 1,066 | 2.67 (1.83, 4.14) |
|  | 216 | 1,895 | 7.13 (5.68, 9.28) | 1,624 | 3.37 (2.22, 5.64) |
|  | 288 | 1,618 | 8.07 (6.43, 10.61) | 1,100 | 5.70 (4.32, 7.80) |
| LMI ^c^ | 120 | 2,932 | 12.07 (11.5, 12.69) | 2,880 | 12.98 (12.43, 13.55) |
|  | 144 | 2,862 | 12.64 (11.95, 13.42) | 2,750 | 13.28 (12.63, 14.00) |
|  | 168 | 2,406 | 13.39 (12.70, 14.10) | 2,353 | 14.87 (13.90, 15.92) |
|  | 192 | 1,266 | 13.56 (12.81, 14.3) | 1,066 | 15.93 (14.76, 16.98) |
|  | 216 | 1,895 | 13.86 (13.17, 14.64) | 1,624 | 17.19 (16.16, 18.18) |
|  | 288 | 1,618 | 14.8 (13.95, 15.82) | 1,100 | 17.45 (16.25, 18.83) |
| BMD ^c^ | 120 | 2,965 | 0.77 (0.74, 0.81) | 2,900 | 0.78 (0.75, 0.82) |
|  | 144 | 2,865 | 0.85 (0.80, 0.90) | 2,756 | 0.84 (0.80, 0.89) |
|  | 168 | 2,406 | 0.96 (0.91, 1.01) | 2,354 | 0.95 (0.90, 1.01) |
|  | 192 | 2,050 | 1.00 (0.96, 1.05) | 1,952 | 1.06 (0.99, 1.12) |
|  | 216 | 1,902 | 1.04 (0.99, 1.09) | 1,634 | 1.14 (1.08, 1.21) |
|  | 288 | 1,619 | 1.19 (1.13, 1.26) | 1,101 | 1.32 (1.23, 1.39) |
| Weight (in kilograms) ^d^ | 120 | 3,105 | 33.60 (29.6, 38.8) | 3,045 | 33 (29.40, 37.80) |
|  | 144 | 2,904 | 42.80 (37.00, 50.20) | 2,801 | 40.60 (35.80, 47.60) |
|  | 168 | 2,433 | 53.40 (47.80, 60.20) | 2,394 | 53.00 (46.80, 61.35) |
|  | 192 | 1,599 | 57.00 (52.00, 64.00) | 1,330 | 66.00 (59.00, 74.00) |
|  | 216 | 1,986 | 61.00 (55.10, 68.40) | 1,683 | 70.40 (63.70, 79.00) |
|  | 288 | 1,671 | 64.90 (58.70, 75.55) | 1,131 | 79.00 (70.70, 88.50) |
| Height (in centimeters) ^d^ | 120 | 3,074 | 139.05 (135.0, 143.4) | 3,027 | 139.8 (135.7, 143.9) |
|  | 144 | 2,904 | 151.4 (146.4, 156.2) | 2,797 | 149.8 (145.1, 154.7) |
|  | 168 | 2,438 | 162.1 (157.8, 166.3) | 2,394 | 165.2 (158.9, 170.8) |
|  | 192 | 1,666 | 165.0 (160.0, 170.0) | 1,361 | 178.0 (173.0, 183.0) |
|  | 216 | 1,988 | 165.1 (161.1, 169.2) | 1,683 | 179.0 (174.5, 183.4) |
|  | 288 | 1,672 | 165.8 (161.9, 170.0) | 1,131 | 180.0 (175.5, 184.5) |

IQR = interquartile range; BMI, body mass index (weight in kilograms/height^2^ in meters); FMI, fat mass index (fat mass in kilograms/height^2^ in meters); LMI, lean mass index (lean mass in kilograms/height^2^ in meters); BMD, bone mineral density.

^a^Full description of the Avon Longitudinal Study of Parents and Children (ALSPAC) is described elsewhere; [2–6].

^b^BMI was calculated using objectively measured weight and height that were collected from different sources (i.e., routine clinic visits, information collected from midwives, linkage to child health records) between birth and age 24 years. Information on weight was collected at research clinic visit annually up to the age 14 years and further clinic measurements at the ages 16, 18, and 24 years using the Tanita Body Fat Analyzer (Tanita TBFUK Ltd.) to the nearest 50 grams. During the same clinic visits, height (standing) was measured to the nearest millimeter with shoes and socks removed using a Holtain stadiometer (Holtain Ltd, Crymych, Pembs, UK).

^c^Fat mass, lean mass, and bone mineral density (BMD) were derived using a Lunar Prodigy dual emission x ray absorptiometry (DEXA) scanner (GE Medical Systems Lunar, Madison, WI, USA). Fat mass index (FMI) and lean mass index (LMI) were calculated by dividing each measure (in kilograms) by height^2^ (in meters). The bone mineral density (BMD) is calculated for the whole body excluding the head values.

^d^For uniformity of the analyses, we restricted analyses of weight and height to measurements taken at the same ages FMI, LMI, and BMD were measured.

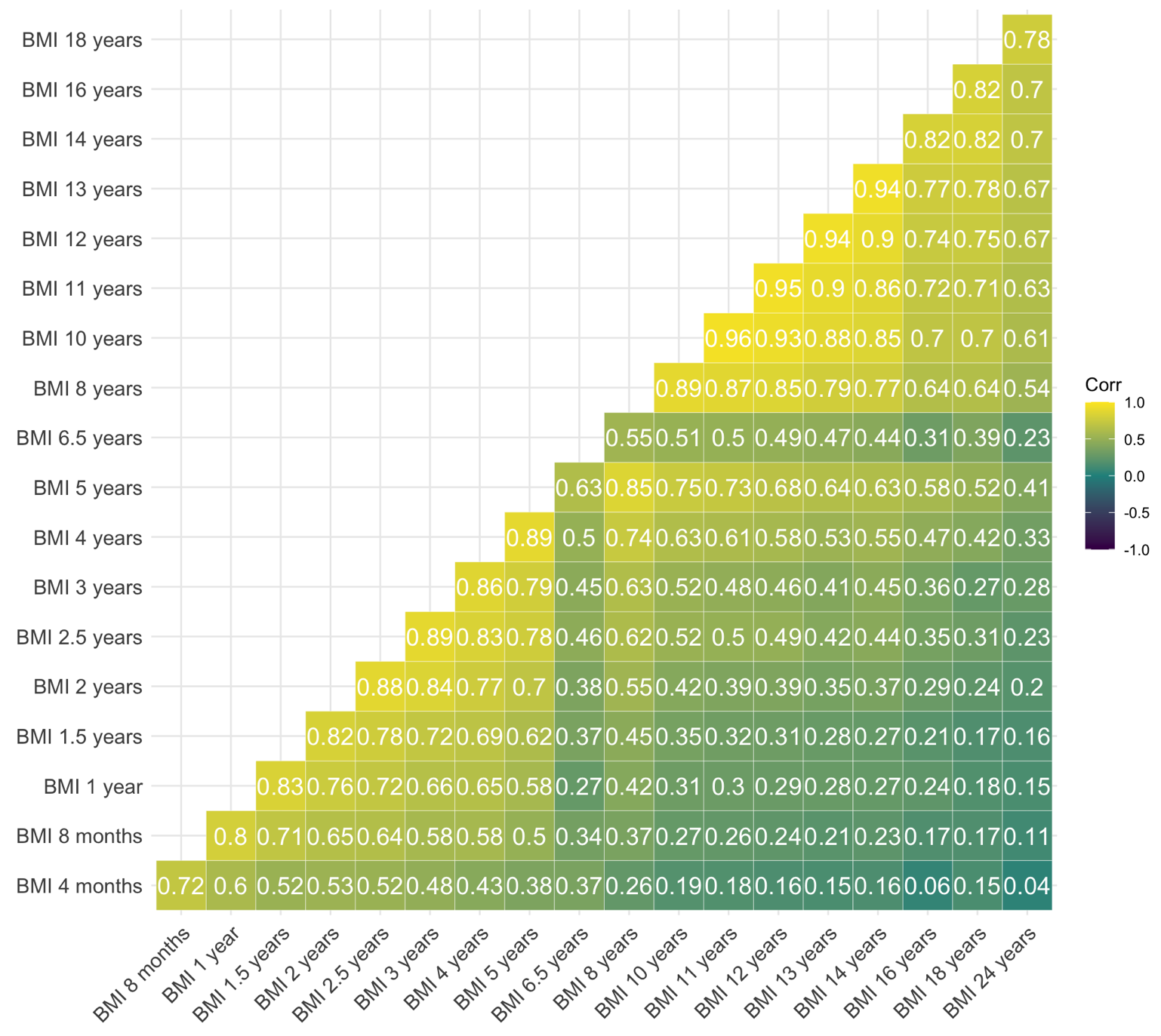

**Figure S1:** Correlation matrix of body mass index (BMI; weight in kilograms/height^2^ in meters) across childhood and adolescence in the Avon Longitudinal Study of Parents and Children (ALSPAC). Numerous measurements on weight and height were collected from different sources (i.e., routine clinic visits, information collected from midwives, linkage to child health records) between birth and age 24 years. Information on weight was collected at research clinic visits annually up to the age 14 years and further clinic measurements at the ages 16, 18, and 24 years using the Tanita Body Fat Analyzer (Tanita TBFUK Ltd.) to the nearest 50 grams. During the same clinic visits, height (standing) was measured to the nearest millimeter with shoes and socks removed using a Holtain stadiometer (Holtain Ltd, Crymych, Pembs, UK). Information on child and adolescent BMI (weight in kilograms / height squared in meters) was derived using weight and height measurement obtained during clinic visits.

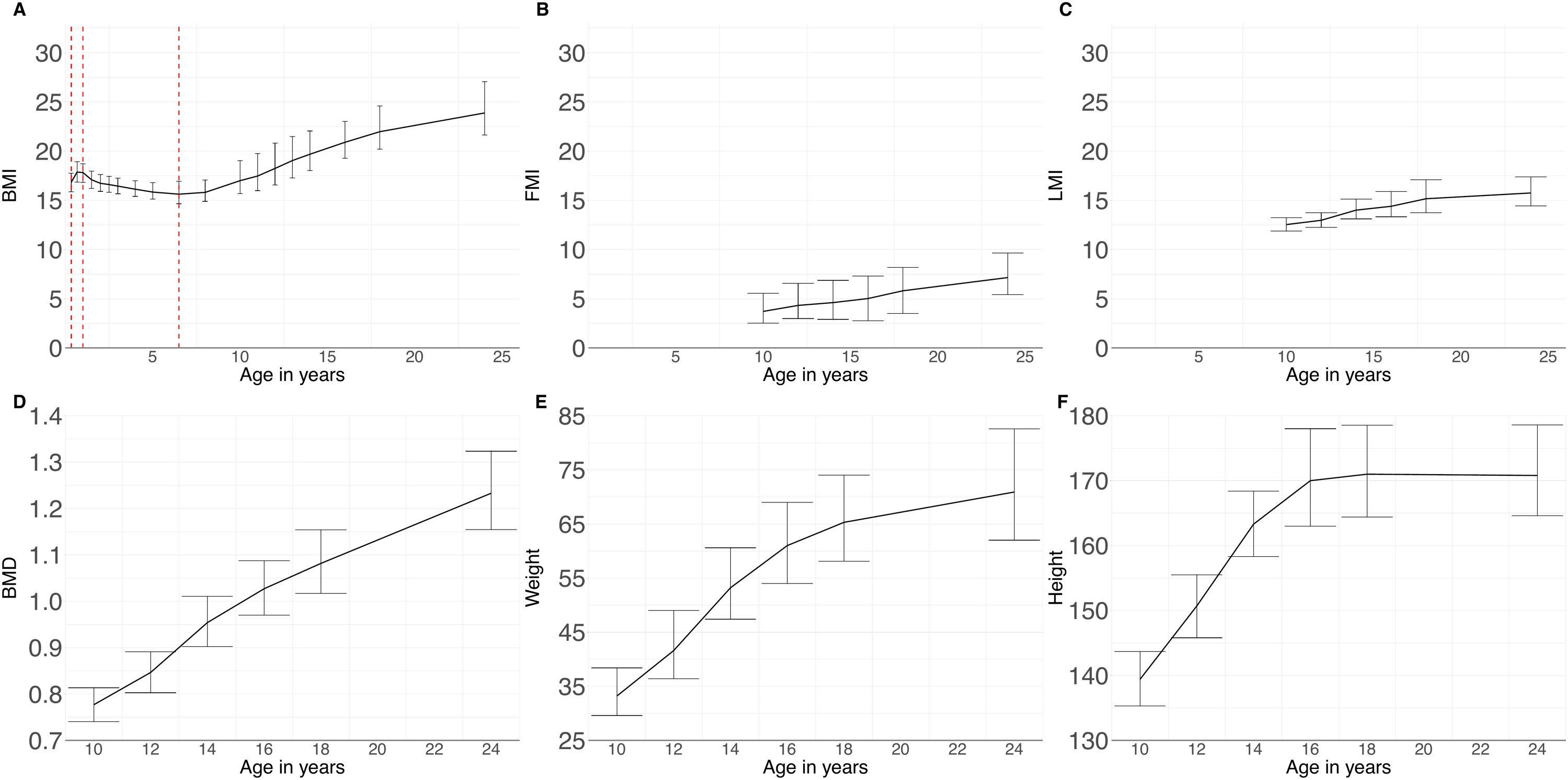

**Figure S2:** **A.** Median of body mass index (BMI; weight in kilograms/height^2^ in meters) trajectories across childhood and adolescence. The BMI trajectory was derived using spline modeling. Prior to deriving the trajectory. For the spline modeling three spline points (knots shown in red) were placed: knot 1, between age 4 months and 1 year; knot 2, between age 1 year and 6.5 years; and knot 3, between age 6.5 year and 24 years. Intercept reflect BMI value at age 4 months. **B-F** Median fat mass index (FMI; fat mass in kilograms/height^2^ in meters), lean mass index (LMI; lean mass in kilograms/height^2^ in meters, bone mineral density (BMD; in gram/cm^2^), weigh, and height trajectories across childhood and adolescents, respectively. FMI, LMI, BMD, weight, and height trajectory parameters were derived using mixed effects models [7]. Error bars represent the interquartile range.

**Table S2:** Sample size of the combined anorexia nervosa and body mass index polygenic scores groups in the Avon Longitudinal Study of Parents and Children^a^

|  | **Female** | **Male** |
| --- | --- | --- |
| **PGS category ^b^** | **N** | **N** |
| Low AN / high BMI | 778 | 702 |
| High AN / high BMI | 137 | 114 |
| High AN / low BMI | 765 | 715 |
| Low AN / low BMI | 2,749 | 2,694 |

^a^ Full description of the Avon Longitudinal Study of Parents and Children is described elsewhere; [2–6].

^b^ Categorical variable derived from dichotomizing the AN-PGS and the BMI-PGS. For both the AN and the BMI PGS, individuals with scores at or greater than the 8^th^ decile point were regarded as the “high PGS group” while those with scores lower were considered “low PGS group”. From the dichotomized AN-PGS and BMI-PGS we were able to create a categorical variable with four levels (1) low AN-PGS / low BMI-PGS (2) high AN-PGS / low BMI-PGS (3) high AN-PGS / high BMI-PGS (4) low AN-PGS / high BMI-PGS. The “low AN-PGS/low BMI-PGS” group was used as the reference category in the extreme analyses.

**Table S3:** Associations of the combined anorexia nervosa and body mass index polygenic score with the body composition measures stratified for biological sex using linear mixed models in the Avon Longitudinal Study of Parents and Children: Post-hoc analyses of the BMI trajectory ^a b^

|  |  | **Female** | | | | **Male** | | | |
| --- | --- | --- | --- | --- | --- | --- | --- | --- | --- |
| **Trajectory** | **Comparison ^b^** | **Beta** | **SE** | **Z ratio** | **P ^c^** | **Beta** | **SE** | **Z ratio** | **P ^c^** |
| **BMI** | Low AN PGS and low BMI PGS - High AN PGS and low BMI PGS | 0.002 | 0.006 | 0.270 | 0.99 | -0.001 | 0.006 | -0.153 | 1 |
|  | Low AN PGS and low BMI PGS - High AN PGS and high BMI PGS | -0.102 | 0.014 | -7.044 | **<0.001*** | -0.098 | 0.013 | -7.775 | **<0.001*** |
|  | Low AN PGS and low BMI PGS - Low AN PGS and high BMI PGS | -0.093 | 0.006 | -14.375 | **<0.001*** | -0.071 | 0.006 | -12.308 | **<0.001*** |
|  | High AN PGS and low BMI PGS - High AN PGS and high BMI PGS | -0.104 | 0.015 | -6.807 | **<0.001*** | -0.097 | 0.013 | -7.315 | **<0.001*** |
|  | High AN PGS and low BMI PGS - Low AN PGS and high BMI PGS | -0.095 | 0.008 | -11.690 | **<0.001*** | -0.071 | 0.007 | -9.708 | **<0.001*** |
|  | High AN PGS and high BMI PGS - Low AN PGS and high BMI PGS | 0.009 | 0.015 | 0.567 | 0.94 | 0.026 | 0.013 | 1.990 | 0.19 |
| **FMI** | Low AN PGS and low BMI PGS - High AN PGS and low BMI PGS | 0.013 | 0.018 | 0.720 | 0.89 | -0.015 | 0.025 | -0.613 | 0.93 |
|  | Low AN PGS and low BMI PGS - High AN PGS and high BMI PGS | -0.274 | 0.042 | -6.583 | **<0.001*** | -0.390 | 0.053 | -7.304 | **<0.001*** |
|  | Low AN PGS and low BMI PGS - Low AN PGS and high BMI PGS | -0.245 | 0.019 | -13.125 | **<0.001*** | -0.323 | 0.025 | -13.135 | **<0.001*** |
|  | High AN PGS and low BMI PGS - High AN PGS and high BMI PGS | -0.287 | 0.044 | -6.563 | **<0.001*** | -0.375 | 0.056 | -6.675 | **<0.001*** |
|  | High AN PGS and low BMI PGS - Low AN PGS and high BMI PGS | -0.258 | 0.023 | -11.100 | **<0.001*** | -0.308 | 0.031 | -10.016 | **<0.001*** |
|  | High AN PGS and high BMI PGS - Low AN PGS and high BMI PGS | 0.029 | 0.044 | 0.654 | 0.91 | 0.067 | 0.056 | 1.194 | 0.63 |
| **LMI** | Low AN PGS and low BMI PGS - High AN PGS and low BMI PGS | 0.006 | 0.003 | 1.605 | 0.38 | 0.000 | 0.003 | -0.008 | **1** |
|  | Low AN PGS and low BMI PGS - High AN PGS and high BMI PGS | -0.027 | 0.008 | -3.415 | **0.0036*** | -0.032 | 0.008 | -4.255 | **<0.001*** |
|  | Low AN PGS and low BMI PGS - Low AN PGS and high BMI PGS | -0.033 | 0.004 | -9.057 | **<0.001*** | -0.027 | 0.003 | -7.892 | **<0.001*** |
|  | High AN PGS and low BMI PGS - High AN PGS and high BMI PGS | -0.033 | 0.008 | -3.913 | **<0.001*** | -0.032 | 0.008 | -4.043 | **<0.001*** |
|  | High AN PGS and low BMI PGS - Low AN PGS and high BMI PGS | -0.038 | 0.004 | -8.514 | **<0.001*** | -0.027 | 0.004 | -6.310 | **<0.001*** |
|  | High AN PGS and high BMI PGS - Low AN PGS and high BMI PGS | -0.005 | 0.008 | -0.631 | 0.92 | 0.005 | 0.008 | 0.602 | 0.93 |
| **BMD** | Low AN PGS and low BMI PGS - High AN PGS and low BMI PGS | 0.007 | 0.003 | 2.037 | 0.17 | -0.003 | 0.003 | -0.771 | 0.87 |
|  | Low AN PGS and low BMI PGS - High AN PGS and high BMI PGS | -0.021 | 0.008 | -2.687 | **0.04*** | -0.018 | 0.007 | -2.492 | 0.06 |
|  | Low AN PGS and low BMI PGS - Low AN PGS and high BMI PGS | -0.029 | 0.003 | -8.308 | **<0.001*** | -0.023 | 0.003 | -6.790 | **<0.001*** |
|  | High AN PGS and low BMI PGS - High AN PGS and high BMI PGS | -0.027 | 0.008 | -3.399 | **0.0038*** | -0.016 | 0.008 | -2.032 | 0.17 |
|  | High AN PGS and low BMI PGS - Low AN PGS and high BMI PGS | -0.036 | 0.004 | -8.243 | **<0.001*** | -0.021 | 0.004 | -4.820 | **<0.001*** |
|  | High AN PGS and high BMI PGS - Low AN PGS and high BMI PGS | -0.008 | 0.008 | -0.997 | 0.75 | -0.005 | 0.008 | -0.603 | 0.93 |
| **Weight** | Low AN PGS and low BMI PGS - High AN PGS and low BMI PGS | 0.008 | 0.008 | 0.923 | 0.79 | 0.008 | 0.008 | 0.923 | 0.79 |
|  | Low AN PGS and low BMI PGS - High AN PGS and high BMI PGS | -0.096 | 0.019 | -4.996 | **<0.001*** | -0.096 | 0.019 | -4.996 | **<0.001*** |
|  | Low AN PGS and low BMI PGS - Low AN PGS and high BMI PGS | -0.107 | 0.009 | -12.340 | **<0.001*** | -0.107 | 0.009 | -12.340 | **<0.001*** |
|  | High AN PGS and low BMI PGS - High AN PGS and high BMI PGS | -0.104 | 0.020 | -5.134 | **<0.001*** | -0.104 | 0.020 | -5.134 | **<0.001*** |
|  | High AN PGS and low BMI PGS - Low AN PGS and high BMI PGS | -0.115 | 0.011 | -10.605 | **<0.001*** | -0.115 | 0.011 | -10.605 | **<0.001*** |
|  | High AN PGS and high BMI PGS - Low AN PGS and high BMI PGS | -0.011 | 0.020 | -0.536 | 0.95 | -0.011 | 0.020 | -0.536 | 0.95 |
| **Height** | Low AN PGS and low BMI PGS - High AN PGS and low BMI PGS | 0.001 | 0.002 | 0.415 | 0.98 | -0.001 | 0.002 | -0.586 | 0.94 |
|  | Low AN PGS and low BMI PGS - High AN PGS and high BMI PGS | 0.002 | 0.004 | 0.560 | 0.94 | 0.002 | 0.005 | 0.519 | 0.95 |
|  | Low AN PGS and low BMI PGS - Low AN PGS and high BMI PGS | -0.002 | 0.002 | -0.754 | 0.87 | -0.006 | 0.002 | -2.819 | **0.03*** |
|  | High AN PGS and low BMI PGS - High AN PGS and high BMI PGS | 0.002 | 0.005 | 0.360 | 0.98 | 0.004 | 0.005 | 0.748 | 0.88 |
|  | High AN PGS and low BMI PGS - Low AN PGS and high BMI PGS | -0.002 | 0.002 | -0.928 | 0.79 | -0.005 | 0.003 | -1.784 | 0.28 |
|  | High AN PGS and high BMI PGS - Low AN PGS and high BMI PGS | -0.004 | 0.005 | -0.851 | 0.83 | -0.009 | 0.005 | -1.717 | 0.32 |

AN = anorexia nervosa; PGS = polygenic score; BMI = body mass index (weight in kilograms/height^2^ in meters); FMI = fat mass index (fat mass in kilogram /height^2^ in meters); LMI = lean mass index (lean mass in kilograms/height^2^ in meters); BMD = bone mineral density (gram/cm^2^).

^a^ Full description of the Avon Longitudinal Study of Parents and Children is described elsewhere; [18–22].

^b^ Categorical variable derived from dichotomizing the AN-PGS and the BMI-PGS. For both the AN and the BMI PGS, individuals with PGS scores at or greater than the 8^th^ decile point were regarded as the “high PGS group” while those with scores lower were considered the “low PGS group”. From the dichotomized AN-PGS and BMI-PGS we were able to create a categorical variable with two levels (1) high AN-PGS and high BMI-PGS (2) high AN-PGS and low BMI-PGS.

^c^ Post-hoc comparisons corrected for multiple testing using Tukey's adjustment.

* Significant after accounting for multiple testing.

**References**

1.	McCarthy, S.; Das, S.; Kretzschmar, W.; Delaneau, O.; Wood, A.R.; Teumer, A.; Kang, H.M.; Fuchsberger, C.; Danecek, P.; Sharp, K.; et al. A reference panel of 64,976 haplotypes for genotype imputation. *Nat. Genet.* **2016**, *48*, 1279–1283.

2.	Boyd, A.; Golding, J.; Macleod, J.; Lawlor, D.A.; Fraser, A.; Henderson, J.; Molloy, L.; Ness, A.; Ring, S.; Smith, G.D. Cohort profile: The ’Children of the 90s’-The index offspring of the avon longitudinal study of parents and children. *Int. J. Epidemiol.* **2013**, *42*, 111–127.

3.	Golding, J. The Avon Longitudinal Study of Parents and Children (ALSPAC)--study design and collaborative opportunities. *Eur. J. Endocrinol.* **2004**, U119–U123.

4.	Golding; Pembrey; Jones; The Alspac Study Team ALSPAC-The Avon Longitudinal Study of Parents and Children. *Paediatr. Perinat. Epidemiol.* **2001**, *15*, 74–87.

5.	Fraser, A.; Macdonald-wallis, C.; Tilling, K.; Boyd, A.; Golding, J.; Davey smith, G.; Henderson, J.; Macleod, J.; Molloy, L.; Ness, A.; et al. Cohort profile: The avon longitudinal study of parents and children: ALSPAC mothers cohort. *Int. J. Epidemiol.* **2013**, *42*, 97–110.

6.	Northstone, K.; Lewcock, M.; Groom, A.; Boyd, A.; Macleod, J.; Timpson, N.; Wells, N. The Avon Longitudinal Study of Parents and Children (ALSPAC): an update on the enrolled sample of index children in 2019. *Wellcome Open Res.* **2019**, *4*, 51.

7.	Herle, M.; Micali, N.; Abdulkadir, M.; Loos, R.; Bryant, R.; Hübel, C.; Bulik, C.M.; Stavola, B.L. De Identifying typical trajectories in longitudinal data : modelling strategies and interpretations. *Eur. J. Epidemiol.* **2020**.
